# Clinical Outcomes, Privacy-Preserving Behavioral Phenotyping, and Automated Risk-Signal Characterization with Digital Journaling: A Randomized Controlled Trial and Naturalistic Study

**DOI:** 10.64898/2026.04.04.26349881

**Authors:** Michael P. Milham, Daniel M. Low, Alp Erkent, Julia Trabulsi, Mirelle Kass, Reinder Vos de Wael, Sailaja Yenepalli, Yanyi Wang, Michael Leyden, Chloe Jordan, Giovanni A. Salum, Lindsay Alexander, Gabriel Schubiner, Lauren Hendrix, Maki S. Koyama, Luke Mears, Roxanne McAdam, Curt White, Kathleen Merikangas, Theodore D. Satterthwaite, Alexandre R. Franco, Arno Klein, Harold Koplewicz, Bennett Leventhal, Michelle Freund, Gregory Kiar

## Abstract

**Background:** Digital journaling may serve both as a low-intensity mental health intervention and as a source of longitudinal behavioral data. Its clinical effects and its potential for privacy-preserving behavioral phenotyping and risk-signal characterization require separate evaluation.

**Objective:** This study evaluated the clinical effects of guided and unguided digital journaling and explored whether longitudinal journaling data could characterize reproducible patterns in mood and classifier-derived distress and risk signals.

**Methods:** Part I was an 8-week randomized controlled trial with a 1-month follow-up among 665 assigned young adults with mild-to-moderate symptoms of anxiety or depression recruited via Prolific Academic. Participants were randomized to guided journaling plus mood check, unguided journaling plus mood check, mood check only, or passive control. Available follow-up PROMIS outcomes from 423 participants contributed to the intention-to-treat models. Part II comprised exploratory, non-preregistered analyses of more than 12,000 self-initiated mood observations and classifier-derived distress and risk signals. Selected behavioral dynamics were examined in an independent general-population dataset of 16,630 users.

**Results:** Of 665 assigned participants, 158 (23.8%) were not invited to follow-up because of incomplete onboarding/linkage or administrative issues; 84 (12.6%) were invited but provided no follow-up outcomes. Overall, 423 (63.6%) provided at least one follow-up outcome, with similar availability across conditions (61.5%–65.1%). Primary contamination-corrected analyses excluded nine passive-control participants who used the study app and yielded small anxiety estimates favoring journaling, with the largest arm-level estimate observed for guided journaling at week 12 (Cohen *d*=0.25). Secondary pooled estimates were approximately d=0.16 at week 8 and d=0.19 at week 12. Neither the arm-level nor pooled comparisons survived correction for multiple comparisons. Complementary Bayesian estimates were directionally consistent with lower anxiety but indicated substantial uncertainty regarding effect magnitude. We found no evidence of benefit for depression or functional impairment. In Part II, classifier-derived high-risk signals were more common among younger users (odds ratio 0.81 per year of age; P=.013). Classifier-derived distress and self-reported energy varied by time of day, and mood and risk states exhibited short-term persistence and escalation patterns. Selected behavioral dynamics were reproduced in the independent general-population dataset, although absolute rates differed between samples.

**Conclusions:** The randomized trial yielded small, uncertain anxiety estimates favoring journaling, but these did not establish clinical benefit as no comparison survived statistical correction for multiple comparisons. No benefit was demonstrated for depression or functional impairment. Separately, the exploratory behavioral analyses showed that privacy-preserving longitudinal journaling data could be used to characterize reproducible temporal patterns in mood and classifier-derived distress and risk signals. These findings support further evaluation of digital journaling as a behavioral measurement and risk-signal characterization platform and motivate future trials of more targeted guided-journaling approaches matched to specific symptoms or therapeutic mechanisms. The study also highlights opportunities and procedural challenges in online panel-based digital trials.

**Trial Registration:** ClinicalTrials.gov NCT07126275.

## INTRODUCTION

Digital mental health mobile apps are used by millions globally for self-monitoring, reflection, and symptom management,[1–3] generating large volumes of real-world behavioral data.[4,5] However, proprietary restrictions and data aggregation render much of this information inaccessible, obscuring clinically relevant dynamics. As a result, much of the empirical study of mental health continues to largely rely on infrequently administered symptom questionnaires,[6] which only capture snapshots of psychological states rather than the dynamic fluctuations in emotions, thoughts, and behaviors that characterize everyday life.

In contrast to traditional research assessments that attempt to summarize mental health status at discrete time points, often separated by months, ecological momentary assessment (EMA) methods using smartphone applications allow higher-frequency characterization of mental and behavioral states in both research and clinical settings.[7,8] These approaches typically use structured prompts delivered at scheduled intervals. At the same time, many consumer-facing digital platforms generate large volumes of behavioral data during natural use. Despite their scale, these data streams are rarely structured or accessible in ways that support longitudinal modeling of affective dynamics. Bridging this gap requires digital platforms designed to balance therapeutic accessibility with measurement precision, user privacy with research utility, and sustained engagement with scientifically interpretable behavioral signals.

Digital journaling represents one promising modality within this context. Reflective writing is accessible, intrinsically engaging, and grounded in decades of expressive writing research that has demonstrated benefits for emotional processing and self-reflection.[9–12] When paired with structured mood check-ins, journaling platforms can generate repeated self-reports that approximate key features of ecological momentary assessment[6,13] while maintaining the flexibility of self-initiated engagement. Such hybrid designs allow emotional experiences to be captured during naturally occurring moments of reflection rather than only in response to externally prompted assessments.

The clinical and public health need for accessible, scalable, self-guided supports remains substantial, particularly among adolescents and young adults who face persistent barriers to care.[14–16] Digital self-reflection tools offer one pathway toward broader access.[17] Assessing their potential clinical value requires reliable estimates of benefit, uncertainty, and participation under the conditions of use. Scalability alone does not establish efficacy. The capacity of digital platforms to support longitudinal behavioral phenotyping and data collection therefore warrants evaluation separately from their therapeutic effects.[18,19]

Scalable engagement is not only a public health goal but also a prerequisite for detecting reproducible emotional patterns at individual and population levels. Sustained, high-frequency data collection creates opportunities to identify temporal patterns that inform early detection, monitoring, and risk stratification.[18,19] Affective science emphasizes variability constructs such as instability, inertia, and volatility as markers of emotional dysregulation, including that seen in anxiety. These dynamics capture general dysregulation, mood persistence, and short-term fluctuations that may be obscured when emotional experience is summarized only by mean symptom levels.[20–22]

Emotional experience is multidimensional.[23,24] In the circumplex model of affect, valence (pleasantness–unpleasantness) and arousal (energy or activation) form orthogonal axes[23] whose joint dynamics may reveal regulatory trajectories hidden by unidimensional mood scales. Abrupt downward shifts, persistence of negative states, and interactions between low pleasantness and elevated energy may constitute informative second-order signals beyond mean symptom levels.[22] Although such models have been widely applied to characterize individuals with mood and anxiety disorders in controlled research settings,[25] there remains a need to understand affective dynamics in broader population samples under natural conditions. Detecting such patterns in large-scale behavioral data requires automated classification methods[26]. Recent advances in natural language processing enable real-time analysis of emotional content and distress signals,[27–29] creating opportunities to augment structured mood reports with behavioral indicators extracted from journaling content.

Digital platforms that combine active mood self-report with AI-assisted classification of journaling content create two complementary behavioral signals: self-reported affect and automated indicators of distress or risk.[30,31] When implemented with strong privacy protections and de-identification procedures, such systems enable aggregation of structured behavioral features (e.g., mood ratings, risk classifications, and engagement metadata) without retaining raw text. This architecture creates two immediate opportunities: user-level support through reflective writing and connection to potential resources, and population-level monitoring through characterization of temporal and demographic patterns. With further validation, such systems may also support remote symptom monitoring that complements episodic clinical encounters in stepped-care settings.[32,33]

The present study evaluated a mobile digital journaling platform (Figure 1) combining structured mood self-reports with automated language-model analysis of free-text journal entries. This approach enables simultaneous capture of subjective affect and algorithmic detection of distress and risk signals. Participants were young adults with anxiety and depressive symptoms (N = 665 assigned, ages 18–30), identified using the GAD-7[34] and PHQ-8[35] and enrolled in an 8-week parallel-arm randomized effectiveness trial with one-month follow-up (ClinicalTrials.gov ID: NCT07126275 [36]). Study arms included guided journaling plus mood checks, unguided journaling plus mood checks, mood checks only, and passive control (who received materials and resources for mental health). Part I assessed whether assignment to journaling conditions was associated with reductions in anxiety and depressive symptoms and improvements in functional impairment, measured using the PROMIS[37] Anxiety and Depression scales and the WHODAS, respectively.[38]

**Figure 1.**
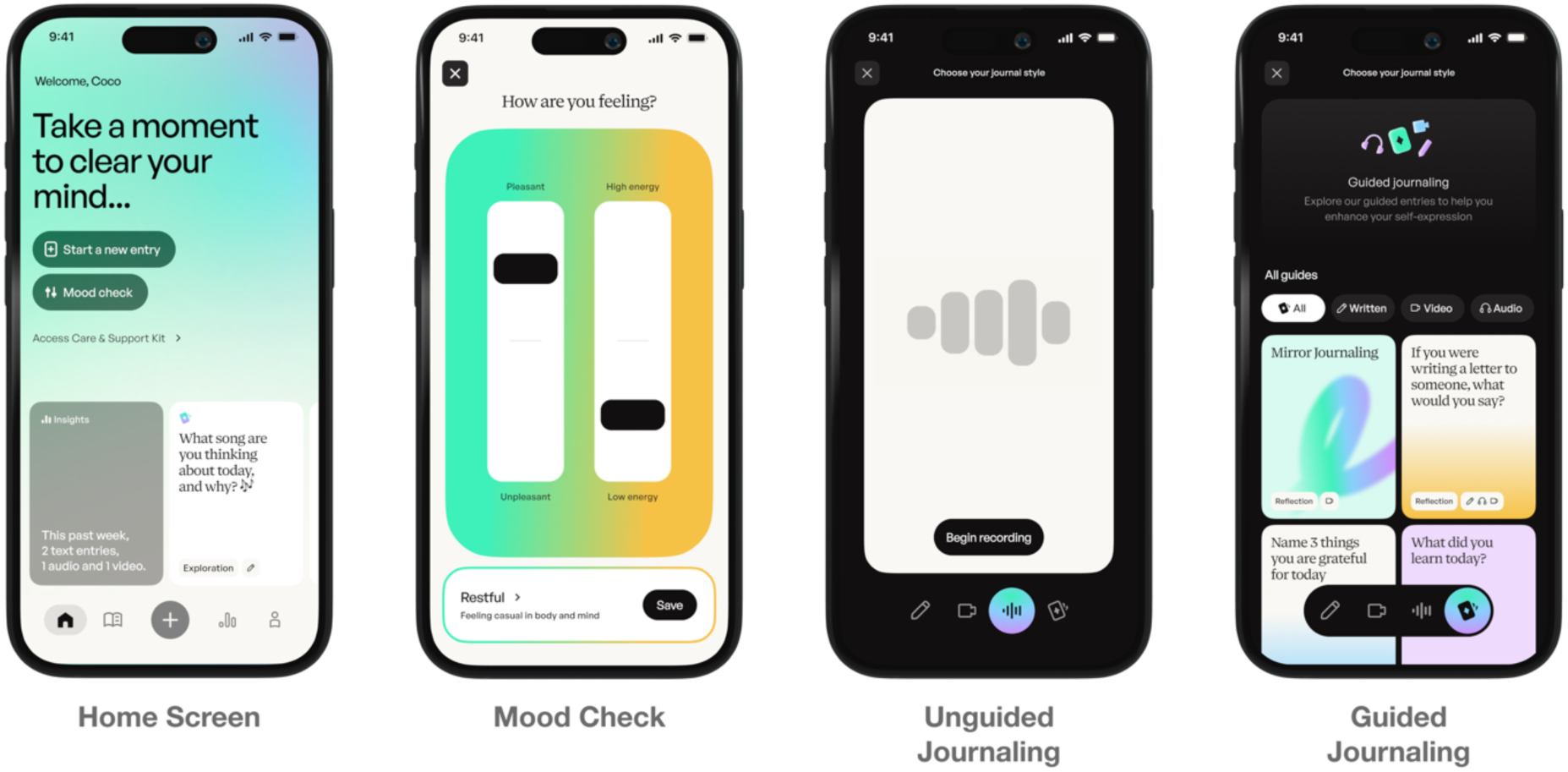
Mirror Journal App. The user interface of the Mirror Journal platform evaluated in the trial. (A) The Home Screen allows users to initiate a new mood check and journal entry. (B) Unguided journaling provides a free-form writing interface, which serves as the source for passive, automated linguistic risk classification, with options for text, video, and audio input. (C) Guided journaling presents reflective prompts for structured writing (text only). (D) The Care and Support Kit provides immediate access to external clinical and crisis resources.

The automated classification framework was designed to identify clinically meaningful signals of distress and potential risk. High-risk classifications required indicators such as imminent intent, planning, or access to lethal means, whereas concerning classifications captured broader indicators of distress or problematic behaviors, including suicidal ideation without imminent planning, recent self-harm, severe distress, high-risk sexual behavior, addiction-related language, or bullying.

Part II leveraged high-frequency longitudinal data from the trial (over 12,000 self-initiated mood-check entries) to derive clinically relevant behavioral signals. Analyses focused on four core areas: (1) population-level and age-related differences in high-risk entry prevalence; (2) temporal structure,[39,40] including time-of-day and weekly variation in self-reported affect (pleasantness and energy) and automated distress signals[41]; (3) short-term transition dynamics reflecting persistence and escalation of mood and risk states; and (4) affective volatility[20–22] (short-term fluctuation across affect dimensions) and its relationship to acute valence drops and trajectories preceding high-risk entries.

As described below, randomized evaluation yielded uncertain between-group anxiety estimates in the direction of the a priori intervention hypothesis, although the frequentist contrasts did not remain significant after correction for multiple comparisons. No comparable effects were observed for depressive symptoms or functional impairment. Complementary Bayesian analyses were used to characterize the direction, magnitude, and uncertainty of the estimated effects rather than as an independent confirmatory test. In parallel, the longitudinal engagement data generated by the trial revealed reproducible temporal structure in affective dynamics and distress signals, including time-of-day variation, state persistence, and short-term escalation patterns. Selected behavioral dynamics were subsequently reproduced in an independent dataset of general population users.

## METHODS

### Overview

The randomized and exploratory components addressed distinct questions. Part I evaluated whether assignment to guided or unguided journaling improved anxiety, depressive symptoms, or functional impairment relative to comparison conditions. Part II comprised exploratory, non-preregistered analyses of longitudinal affect and classifier-derived distress and risk signals, with selected behavioral dynamics examined in an independent general-population dataset. Findings from Part II were interpreted as evidence regarding behavioral measurement and risk-signal characterization rather than clinical efficacy.

### Study design and participants

The clinical component was an 8-week parallel-arm randomized controlled trial (ClinicalTrials.gov ID: NCT07126275) evaluating whether app-based journaling reduces anxiety and depressive symptoms. We recruited participants online through Prolific Academic (https://www.prolific.com/) and screened them for eligibility using self-report. We included participants based on the following criteria: (1) age 18–30 years, (2) U.S. residency, (3) access to a smartphone or internet-enabled device, and (4) mild to moderate anxiety or depressive symptoms, as indicated by PHQ-8 and GAD-7 (scores 5–14). Participants with severe symptoms (scores 15 and above) were excluded due to the online nature of the study, which prevented the provision of intensive clinical care or monitoring and the potential for unintentionally impacting clinical status. Mirror was intended to be an adjunctive intervention; we did not limit anyone based on other interventions. A total of 665 unique participants received a condition assignment during recruitment (March–May 2025). The implemented age range of 18–30 years was approved by the IRB before recruitment; the registry listed ages 18–25 and was not updated to reflect this change. The trial ended after the planned recruitment and follow-up activities were completed; it was not stopped early. All procedures were approved by Advarra Institutional Review Board (https://www.advarra.com/) and electronic informed consent was obtained from all participants. The trial record was first submitted to ClinicalTrials.gov on February 11, 2025, before participant enrollment began, and was first publicly posted on August 17, 2025, after enrollment began, following ClinicalTrials.gov quality-control review. The full study protocol is available from the corresponding author upon reasonable request.

### Randomization and allocation implementation

Eligible participants completed consent and baseline assessments through REDCap. Allocation slots within baseline journaling-frequency categories were ordered using two successive random-number sorts. Assignment was ordinarily triggered automatically upon submission of the journaling-frequency response; staff could not skip assignments or select conditions. Additional slots were appended during recruitment through the REDCap administrator, without altering existing assignments or reassigning participants. Upcoming assignments were visible to study personnel. Exceptions involving manually triggered assignments are described under Participant flow.

### Interventions

The intervention was delivered through the Mirror Journal mobile application (https://www.mirrorjournal.com/), a privacy-preserving digital journaling platform developed by the Child Mind Institute. Mirror Journal is part of a broader portfolio of scalable digital mental health tools developed through the Child Mind Institute’s Next Generation Digital Therapeutics (NGDT) program, including initiatives conducted in collaboration with the California Department of Health Care Services (DHCS) through the Children and Youth Behavioral Health Initiative (CYBHI) (note: DHCS and the CYBHI had no role in study design, data collection, analysis, or interpretation of this data).

Participants were randomized to one of four parallel 8-week conditions (Figure 2):

1. *Mood check + guided journaling* (Group 1): Participants first selected their mood and then submitted a journal entry in response to a reflective prompt.
2. *Mood check + unguided journaling* (Group 2): Participants first selected their mood and then engaged in unrestricted free-form journaling.
3. *Mood check only* (Group 3): Participants only selected their mood.
4. *Passive control* (Group 4): Participants did not journal or record mood entries.

**Figure 2.**
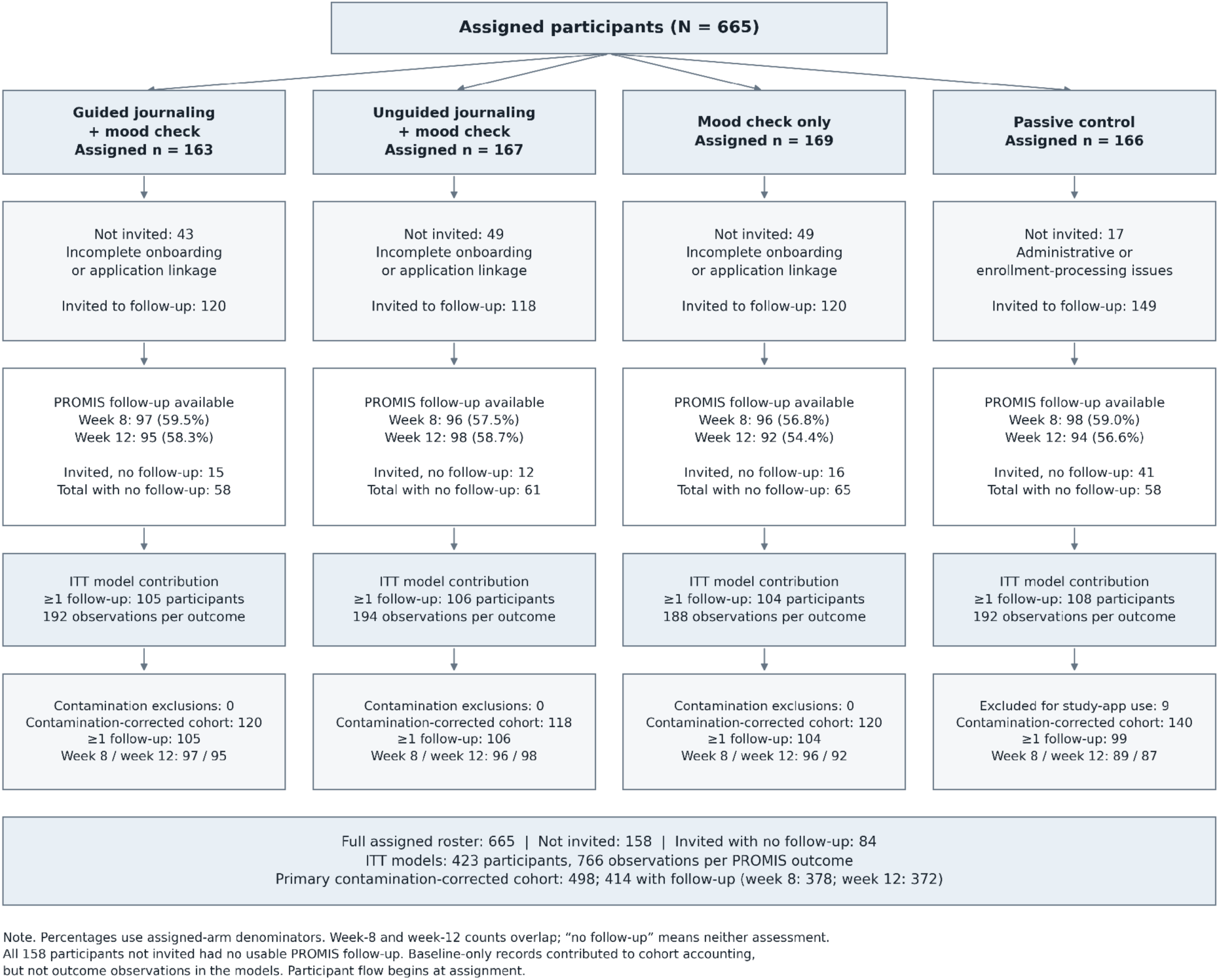
**Participant flow** from condition assignment through follow-up in the 8-week randomized trial (665 assigned participants). Counts distinguish the original extraction, available PROMIS follow-up outcomes, and the retained contamination-corrected analyses. Follow-up proportions use the full assigned denominator. Week-8 and week-12 counts can overlap; participants could return at week 12 after missing week 8. Absence of a usable outcome does not establish why it is missing.

### Masking and participant support

Participants were aware of their assigned activities during intervention delivery and could not be blinded to condition. The intervention was delivered by the application and clinical outcomes were collected through remote self-report; no therapist delivered the journaling intervention and no clinician rated the primary outcomes. Study personnel were not masked to allocation and contacted participants through reminders and technical troubleshooting. Analysts were not masked. Supplementary Methods describes the reminder schedule and operational changes.

Participants in Groups 1–3 were encouraged to engage daily, although this was not required. Assessment compensation was independent of intervention arm and engagement frequency. The assessment schedule for all four groups included screening and baseline assessment, eight weekly surveys, a post-study assessment (week 8), and a one-month follow-up assessment (week 12); completion varied across participants.

Follow-up invitations were sent only to the 507 participants classified as having completed onboarding or study linkage, using the same invitation and reminder procedures across conditions.

Assessments occurred at baseline, weekly during the intervention, post-study (week 8), and follow-up (week 12). Mood check and journaling arms generated longitudinal affect and engagement data.

### Compensation

Participants received fixed payments through Prolific Academic for assessment completion ($3 each for screening, enrollment, and 8 weekly check-ins; $5 each for post-study and follow-up questionnaires; $40 maximum), independent of intervention arm or engagement frequency. Additional compensation reflected task burden: journaling arms (Groups 1-2) received $1.50/entry (maximum $84 over 56 days); mood check only (Group 3) received $0.50/entry (maximum $28); passive control (Group 4) received no additional compensation. Daily entry payments were capped at once daily to prevent artificial inflation of engagement patterns. This compensation structure was approved by the institutional review board.

### Adverse-event ascertainment

Adverse events and serious adverse events were not systematically solicited through a dedicated questionnaire. Participants could report concerns or events to the study team through Prolific Academic’s messaging system and the contact information provided in the consent materials. Available participant communications, withdrawal records, and study documentation were reviewed for reported adverse events and serious adverse events. Automated classifications of journal entries as concerning or high risk were handled through the platform’s safety-response pathway and were not considered adverse events solely on the basis of classifier output.

### Part I. Clinical Outcomes Evaluation

#### Outcome measures

In accord with the trial registration (ClinicalTrials.gov ID: NCT07126275), primary outcomes were PROMIS Anxiety[37] and PROMIS Depression[37] T-scores assessed at baseline, week 8, and week 12 follow-up. Functional impairment measured with WHODAS 2.0[38] served as a secondary outcome. The registry specified outcome measures and assessment timepoints but did not enumerate detailed analytic hypotheses. Post-study questionnaires also collected usability and experience measures (the Mirror Feedback Survey, the MAUQ, the System Usability Scale, and study-and group-specific items); these measures are not analyzed in the present report. No formal qualitative interviews or qualitative analysis were conducted.

#### Study hypotheses and analysis framework

The directional intervention hypothesis that journaling would reduce anxiety and depressive symptoms was specified a priori, and the clinical outcomes and assessment timepoints were specified in the trial registry. The registry did not enumerate the exact analytic contrasts, tailing convention, or multiplicity procedure. Because the intervention was explicitly designed to promote wellbeing and reduce symptoms, the hypothesized direction of the intervention effect was improvement rather than simply any difference between conditions. The four-arm randomized analysis was treated as the primary clinical analysis; pooled journaling-versus-non-journaling and guided-versus-unguided comparisons were secondary. Part II behavioral phenotyping analyses were exploratory and were not preregistered. Analyses in the independent general-population dataset were conducted as replication analyses to assess whether key behavioral patterns reproduced in a distinct sample.

#### Sample size

Enrollment was determined by available recruitment resources and the sampling frame; no formal power analysis was specified in the trial registration. No formal interim efficacy analyses or statistical stopping rules were specified or conducted. A retrospective sensitivity calculation using observed baseline–week-12 complete pairs yielded approximate minimum detectable effects of d=.29–.30 for arm-level comparisons with passive control and d=.21 for the pooled contrast. These calculations did not determine enrollment (Supplementary Methods).

#### Baseline comparisons and follow-up availability

Baseline characteristics were compared between participants invited and not invited to follow-up and, among those invited, between participants with and without at least one follow-up assessment. Continuous variables were compared using Welch’s two-sample *t* test, and categorical variables using Pearson’s chi-square test. Benjamini–Hochberg false discovery rate correction was applied separately within each of the two cohort comparisons across nine baseline characteristics: age, education, PROMIS Anxiety and Depression T-scores, GAD-7, PHQ-8, WHODAS, gender, and race and ethnicity. Each categorical characteristic contributed a single omnibus test to the correction family rather than separate tests for its individual categories. Among participants invited to follow-up, availability of at least one follow-up assessment was compared across assigned conditions using a separate Pearson’s chi-square test.

#### Statistical Analysis

Symptom trajectories were evaluated using baseline-adjusted linear mixed-effects models with random intercepts for participants and fixed effects for condition, follow-up time (week 8, week 12), and their interaction, including baseline symptom scores as covariates. Models were fitted using restricted maximum likelihood. Baseline was a covariate rather than an outcome occasion. Baseline journaling frequency, which informed allocation, was not included as a covariate in these models. The primary analysis compared all four randomized arms simultaneously. Follow-up analyses examined (a) a collapsed comparison of journaling (guided + unguided) versus non-journaling (mood tracking only + passive control), and (b) guided versus unguided journaling.

Missing follow-up outcomes were not imputed. Mixed-effects models used all available follow-up observations under a missing-at-random (MAR) assumption conditional on observed model covariates. Participants with baseline data only contributed to cohort accounting but no outcome observations to these models. Reconciliation of the full assignment roster does not recover missing outcomes or remove the potential for selection bias. The MAR assumption cannot be verified from the observed data.

Analytic populations. The originally extracted cohort comprised 507 of the 665 assigned participants. The reported primary contamination-corrected analyses used 498 members of that extracted cohort after excluding nine passive-control participants who accessed active intervention features. These analyses condition on post-assignment behavior and are not full intention-to-treat (ITT) analyses. ITT sensitivity analyses used the full roster of 665 participants in their originally assigned conditions, without exclusions for onboarding, linkage, app use, or contamination; available PROMIS follow-up outcomes from 423 participants contributed to model estimation. Additional sensitivity analyses refitted the ITT models with years of education, race and ethnicity, and both as covariates, retaining the original model specification otherwise. Finally, active-comparator sensitivity analyses compared journaling conditions with mood checking alone, holding onboarding requirements constant to assess whether the findings persisted under comparable participation demands.

Because the study specified directional hypotheses for the effects of journaling on anxiety and depression, one-tailed tests (α = 0.05) were applied to contrasts comparing journaling versus non-journaling conditions. The mood tracking-only versus passive-control contrast, for which there was no corresponding directional journaling hypothesis, was evaluated using a two-tailed test. For transparency and to assess sensitivity to the tailing convention, corresponding two-tailed P values are also reported for the journaling contrasts. Benjamini–Hochberg false discovery rate (FDR) correction was applied separately within each outcome across all six arm-level contrasts (three intervention arms × two follow-up assessments). For the secondary pooled comparison, correction was applied separately within each outcome across the two follow-up contrasts.

Complementary Bayesian mixed effects ANCOVA models (same model specification as the frequentist models) were also estimated to quantify directional evidence for symptom change and characterize uncertainty in effect estimates. These models describe the same clinical observations and do not provide independent confirmation of efficacy. For each outcome, two models were fitted independently: a four-arm model with passive control as the reference condition, and a pooled model in which guided and unguided journaling were combined into a single journaling condition and mood tracking and passive control into a single non-journaling condition, with non-journaling as the reference. Weakly informative priors were specified based on the PROMIS T-score scale (population mean = 50, SD = 10): Intercept ∼ Normal(50, 20); baseline regression coefficient ∼ Normal(0.5, 0.3); group, timepoint, and interaction coefficients ∼ Normal(0, 10); residual SD ∼ HalfNormal(10). The participant random intercept was assigned the default weakly informative prior of the bambi package, Normal(0, sd) with sd ∼ HalfNormal(s), where s is set automatically by the package from the dispersion of the outcome and the scales of the common-effect priors. Analyses were implemented in Python using the bambi and pymc libraries. Posterior distributions were estimated using the No-U-Turn Sampler (NUTS; target acceptance rate = 0.90) with 4 independent Markov chains, each running 2,000 tuning steps followed by 2,000 sampling draws (8,000 total post-warmup posterior draws). Convergence was assessed using the R-hat statistic; all parameters achieved R-hat ≤ 1.01, and effective sample sizes exceeded 400 for all parameters. We report the posterior mean difference from the reference condition at each timepoint, along with the 90% Highest Density Interval (HDI). A 90% HDI is reported throughout for consistency with the directional framing of the intervention hypotheses. Consistent with the tailing conventions used in the frequentist analyses, the journaling contrasts were evaluated directionally and we report the posterior probability of symptom reduction, P(effect < 0); the mood tracking-only contrast, for which no directional journaling hypothesis applied, was evaluated non-directionally and we report the probability of direction, that is, the posterior probability mass on the larger side of zero. These probabilities are descriptive summaries rather than independent tests: under diffuse priors the directional probability corresponds closely to the one-sided frequentist P value and does not constitute separate evidence. Interpretive weight is therefore placed on posterior magnitude and uncertainty, including the proportion of posterior mass within a region of practical equivalence (ROPE; ±1 T-score point).

### Part II. Measurement of Affect and Risk Dynamics

#### Data source and observations

Mood check data was collected from all active intervention arms (guided journaling, unguided journaling, and mood check only). Risk detection data, however, was only collected from the unguided journaling arm, in line with the app’s design at the time. Each observation was standardized into a user × day × hour time bin, with affect ratings computed as the mean of all mood check-ins within that hour. Entries without valid mood check data were excluded. All timestamps were analyzed in stored local time to preserve time-of-day alignment.

Across the trial period, approximately 12,553 user × day × hour observations were contributed by 357 active users, representing 71.5% of the 499 participants assigned to active conditions. Of the 499 participants assigned to active conditions, 141 did not complete onboarding or application linkage and therefore had no linked behavioral data available for analysis. For analyses requiring entry-level or user-level summaries (e.g., volatility, transitions, and escalation analyses), the corresponding measures were computed from the underlying mood check or journal-entry records.

#### Replication in a general-population sample

To assess generalizability, we replicated all Part II analyses using data from general population users of the Mirror app (ages 18–30, United States) collected between August 1, 2025 and December 31, 2025 (N=16,630 users). In contrast to the clinical trial sample, this dataset reflects routine app use without protocol constraints. Key differences include: (1) no exclusion criteria (the trial excluded individuals with current suicidal ideation); (2) voluntary engagement without financial incentives; and, (3) heterogeneous patterns of use without predefined participation windows. As a result, the dataset captures a broader range of symptom severity, user motivation, and temporal engagement patterns than the trial sample.

All statistical procedures, variable definitions, and privacy protections applied were identical to those used in the trial analyses. Convergent findings across the trial and general-population datasets support the reproducibility of selected affective dynamics and risk-signal patterns across these two engagement contexts. Conversely, any divergence was interpreted as suggesting that these dynamics may vary across engagement contexts and population characteristics.

Secondary research use of the de-identified general-population app data was reviewed as exempt under 45 CFR 46.104(d)(4). Consistent with the privacy policy agreed to at registration, only de-identified behavioral and demographic features were used; investigators had no access to personally identifying information or raw journal text.

#### Privacy

In Mirror Journal, entries are deleted immediately following automated processing (within seconds to minutes). Retention of the raw journal content for research requires explicit opt-in consent, which was not requested for this study. Since participants were recruited via Prolific and data were provided in de-identified form, investigators had no access to personally identifiable information. Consistent with IRB-approved consent procedures, retained data were limited to mood ratings, risk classifications, timestamps, word counts, and engagement metadata; the raw text was not stored.

#### Affect measures

Participants in Groups 1–3 recorded mood using two orthogonal slider bars, ranging from −5 to +5 (excluding the neutral midpoint 0), for a total of 10 selectable points. This process included the option to select mood-related terms from a grid to refine their choices. Pleasantness (P) indexed valence (emotional negativity to positivity), and energy (E) indexed subjective activation. For participants in the journaling arms, the mood check preceded the journal entry. Additionally, a ’vibe check’ was included following journaling to assess immediate post-session mood improvement on a 7-item Likert scale (checking if the entry made them feel better or worse).

### AI-Based Classification of Distress and Risk

Multiple natural language processing approaches can be taken to detect suicidal and other concerning language from text data; a key decision is whether models are deterministic (e.g., BERT-style models that return the same output given the same input) and/or general-purpose and flexible enough to discover issues for which the model was not explicitly trained to detect (e.g., generative LLMs)[27–29].

#### Model Development

Mirro Journal used three complementary language-model components to identify distress and potential safety risk in unguided journal entries: a fine-tuned three-class RoBERTa model, LlamaGuard^45^, and a clinician-designed prompt implemented using Llama 3.2:3B.

The RoBERTa classifier was developed in two stages. Initial training used 120,000 balanced binary-labeled entries from the ROBIN dataset[41] on online forum posts to distinguish suicide- and self-harm–related risk from no-risk content. A second phase of fine-tuning used 2,142 new entries spanning the three final classification categories: no concern (1,112), concerning entries (513), and high-risk entries (517). These included 859 entries from a Mirror pilot study, 533 entries from the Clinician Review dataset, and 750 synthetic entries.

LlamaGuard served as an additional detector of high-risk suicide- and self-harm–related language. Finally, the prompted Llama model was designed to identify a broader range of emotional distress and emerging safety concerns, including domains not emphasized in the suicide-focused RoBERTa training data, such as potential harm to others and sexual violence.

Hierarchical Classification Rule. The three model components were combined using a prespecified hierarchical decision rule. An entry was classified as high risk when either LlamaGuard or RoBERTa assigned a high-risk classification. Among entries not classified as high risk, an entry was classified as concerning when either the prompted Llama model or RoBERTa assigned a concerning classification. All remaining entries were classified as no concern.

This OR-based ensemble was designed to prioritize sensitivity for potentially acute risk while retaining a separate pathway for detecting broader emotional distress and emerging concerns. High-risk classifications took precedence over concerning classifications.

At the time of the study, automated language-model classification was consistently applied to unguided journaling entries and to a limited subset of guided entries. Raw journal text was deleted immediately after automated processing. Retained research data were limited to derived risk classifications, mood ratings, timestamps, word counts, and engagement metadata.

### Component-Model Evaluation

For the clinician review dataset, six clinicians annotated 1,000 ROBIN entries, with each clinician reviewing 400 entries and each entry receiving at least two independent ratings. Clinicians assigned each entry to one of three categories: no concern, concerning, or high risk. Of these entries, 533 were included in the second-stage RoBERTa fine-tuning dataset and 467 were reserved as a held-out test set. The component-model evaluation reported here included the 210 held-out entries for which complete clinician reference labels and predictions from all three model components were available. Synthetic entries were used only for fine-tuning and were not included in the evaluation set.

When clinician ratings differed, the reference label was defined as the highest-severity rating assigned by any reviewer. For example, an entry rated no concern by one reviewer and concerning by another was assigned a concerning reference label. This procedure produced a conservative, safety-oriented reference standard rather than a consensus label.

Each model was evaluated against the reference category corresponding to its intended function. LlamaGuard was evaluated for identification of high-risk entries, the prompted Llama model was evaluated for identification of concerning entries, and the three-class RoBERTa model was evaluated separately for both high-risk and concerning classifications. Sensitivity, specificity, precision, and F1 scores were calculated using one-versus-rest comparisons.

Detailed component-level performance metrics, qualitative false-positive analyses, and performance of the complete hierarchical ensemble are reported in the Supplementary Results. These metrics characterize the individual component models rather than the complete hierarchical ensemble, whose operating characteristics also depend on overlap among model predictions. Automated categories such as these should be interpreted as operational proxies for distress and safety-relevant language rather than as clinical diagnoses or validated measures of underlying clinical constructs.

#### Safety response pathway for distress and risk

When a user either (i) self-reported low pleasantness and/or low energy during a mood check or (ii) submitted a journal entry to which automated risk classification was applied and that was classified as concerning or high risk, the user was presented with a message directing them to the Care and Support Kit, which provides immediate access to external mental health and crisis-support resources. Automated classification did not itself trigger emergency-service contact, clinician outreach, or other involuntary intervention. This approach was designed to make support readily accessible while recognizing the imperfect operating characteristics of automated classification and preserving user privacy.

#### Population-level risk and age-related differences

To examine age-related differences in risk prevalence, we modeled the probability that a journal entry was classified as high risk as a function of user age. Journal entries were first aggregated at the user level to obtain, for each participant, the total number of entries and the number classified as high risk. A binomial generalized linear model with a logit link was then fit using the number of high-risk entries as successes and the remaining entries as failures in a binomial likelihood. Age was included as a continuous predictor.

This formulation preserves information about entry frequency while representing each participant as one aggregated binomial observation. Model coefficients represent changes in the odds that a journal entry is classified as high risk as a function of user age.

To visualize the relationship between age and high-risk probability, predicted probabilities were generated across the observed age range. Uncertainty in the predicted curve was estimated using nonparametric bootstrap resampling of users (1,000 iterations), with percentile-based 95% confidence intervals derived from the bootstrap distribution.

#### Temporal structure

Hours were grouped into eight contiguous 3-hour bins relative to the user’s timezone (beginning with 00:00–03:00) and four 6-hour periods (night, morning, afternoon, evening). Day of week was treated as a seven-level categorical factor. Sampling density varied substantially across the day, with evening windows (18:00–24:00) accounting approximately half of all observations and early-morning windows (03:00–06:00) representing a small minority; caution was applied when interpreting sparsely sampled bins.

#### Transition probability and state persistence

Mood states were assigned to one of four circumplex quadrants based on the joint sign of Pleasantness (P) and Energy (E) slider ratings, each ranging from −5 to +5, with the neutral midpoint (0) excluded from categorization. Pleasantness indexed valence (unpleasant to pleasant), and Energy indexed subjective activation (low to high energy). For classification, scores from +1 to +5 were defined as positive and scores from −1 to −5 as negative. The four quadrants were defined as: (i) Happy/Alert (P positive, E positive), (ii) Anxious/Agitated (P negative, E positive), (iii) Sad/Withdrawn (P negative, E negative), and (iv) Calm/Relaxed (P positive, E negative).

Transition probability matrices were computed using row-normalized cross-tabulations of current versus subsequent states, yielding conditional probabilities of remaining in or transitioning between categories at the next entry. State persistence estimates and 95% confidence intervals were derived using user-clustered bootstrap resampling, with persistence rates first calculated at the user level and then resampled across users. Analyses were conducted separately for mood quadrants and risk states. Because automated risk classification was only consistently available in the unguided journaling condition, risk-state analyses were restricted to that arm. Risk states were assigned only to observations with evidence that automated classification had occurred, defined as either the presence of a journal entry or a positive Concerning or High-Risk classifier output. Observations without journal content or classifier output were treated as unscored rather than classified as Stable. Sensitivity analyses reported in the Supplementary Materials repeated the transition analyses using maximum inter-entry intervals of ≤24 and ≤72 hours, in addition to the primary ≤48-hour specification.

#### Affective volatility profiling and acute affective drop identification

Users with fewer than 10 valid observations were excluded from volatility analyses to avoid unstable estimates of within-person variability based on extremely sparse time series. When multiple entries occurred within the same calendar day, daily mean values were computed and used for drop detection; volatility was calculated from individual observations.

User-level affective volatility was computed separately for pleasantness and energy as the within-user standard deviation across observations[20–22]. To isolate instability independent of baseline affective tone, volatility metrics were residualized with respect to individual mean levels using linear regression.

Acute affective drop events were identified using a forward-window approach. Drops were calculated from the first daily mean to the minimum within a window of four successive observed dates. Analyses evaluated thresholds ranging from ≥2 to ≥5 points, with ≥3 points designated as the primary threshold and used for data visualization. Windows advanced by three observed dates, so adjacent windows shared an endpoint; elapsed calendar time could vary.

For each user, drop susceptibility was quantified as the rate of qualifying drops, defined as the proportion of evaluable windows containing a drop. Associations between affective volatility and drop susceptibility were quantified using Pearson correlations. To examine dimensional specificity, correlations were computed separately between residualized pleasantness volatility and drop rates, and between residualized energy volatility and drop rates. Sensitivity analyses evaluated how these associations varied across different drop thresholds.

To assess whether the volatility–drop associations were robust to data-sufficiency requirements and whether they reflected temporal ordering beyond marginal affective variability, we conducted supplementary sensitivity analyses. We first imposed the additional requirement that users contribute at least five evaluable windows to test whether the observed associations were robust to excluding participants with insufficient longitudinal data to estimate drop susceptibility reliably. We then examined robustness across minimum-observation thresholds and used within-user unrestricted and block-preserving temporal permutations to test whether the observed associations exceeded those expected from marginal affective variability alone.

#### Dissociation between affect volatility and clinical risk

To evaluate whether affective volatility predicted escalation into severity-based risk classifications, volatility metrics were examined in relation to Concerning and High-Risk entries within escalation models. Logistic regression models with user-clustered robust standard errors estimated whether volatility independently was associated with subsequent risk status. Volatility metrics were entered separately from mean affective levels to distinguish short-term fluctuation from sustained low affect.

#### Risk escalation trajectories

To evaluate whether prior risk states predicted near-term escalation, history indicators were constructed for each entry within a user’s time series. Three history definitions were evaluated: (1) presence of at least one high-risk classification within the prior three entries, (2) presence of a high-risk classification within the prior 72 hours, and (3) any prior high-risk classification occurring earlier in the user’s observation history.

For each entry, the outcome of interest was the classification of the subsequent entry, specifically whether the next entry was categorized as high risk or concerning. Entries without a subsequent observation were excluded.

Outcome probabilities were estimated using a user-level rate estimator. For each history condition, the probability of the outcome was first computed separately for each user and then averaged across users, thereby weighting each participant equally regardless of the number of observations contributed. 95% confidence intervals were obtained using bootstrap resampling of users (2,000 iterations).

Analyses were conducted separately for two outcomes: (1) probability that the next entry was classified as high risk and (2) probability that the next entry was classified as concerning. For interpretive context, overall baseline probabilities of each outcome were also computed across the dataset.

#### Software and ethics

Analyses and visualizations were conducted in Python using pandas, numpy, scipy, statsmodels, matplotlib, and seaborn. Modular scripts implemented transition construction, acute affective drop detection, volatility profiling, mixed-effects and GEE modeling, stratified bootstrap estimation, and visualizations. The analysis approach was determined by the investigators; however, data processing scripts were generated iteratively using large language models (Claude, Gemini, and ChatGPT) to assist with code drafting and debugging. Claude and Gemini were primarily used for initial script generation, and ChatGPT was used for independent review and correction. All outputs were manually inspected, tested, and validated.

The completed CONSORT-EHEALTH checklist is provided in Multimedia Appendix 1.

## RESULTS

### Part I. Clinical Outcomes

#### Participants and retention

A total of 665 participants were assigned to guided journaling (n=163), unguided journaling (n=167), mood check only (n=169), or passive control (n=166). Outcome assessments were available for 387 participants (58.2%) at week 8 and 379 (57.0%) at week 12; 423 (63.6%) contributed at least one follow-up assessment. Week-12 availability was similar across conditions: 58.3%, 58.7%, 54.4%, and 56.6%, respectively.

Reasons for missing follow-up differed across conditions. In the app conditions, 141 of 184 participants without any follow-up assessment (76.6%) had not completed onboarding or linkage and were therefore not invited to follow-up assessments. In passive control, 17 of 58 participants without follow-up (29.3%) were not invited because of administrative or enrollment-processing issues, including 15 whose control assignments were manually triggered but who were not notified after the control recruitment quota had been reached. The remaining 41 controls (70.7%) were invited but provided no follow-up assessment. Arm-specific participant flow is shown in Figure 2.

#### Participant demographics and analytic cohort

Baseline demographic and clinical characteristics of all 665 assigned participants are presented in Table S2. The primary contamination-corrected cohort included 498 participants, of whom 414 contributed at least one PROMIS follow-up outcome. Week-8/week-12 outcome counts were 97/95 for guided journaling, 96/98 for unguided journaling, 96/92 for mood check only, and 89/87 for passive control.

#### Adverse events

No adverse events or serious adverse events were reported to the study team during the trial.

#### Baseline characteristics and follow-up availability

Participants invited to follow-up (n=507) and those not invited (due to incomplete onboarding/linkage or administrative issues; n=158) showed no statistically significant differences in age or baseline anxiety and depressive symptoms after false discovery rate correction. Participants not invited reported slightly more education (15.47 vs 15.04 years; adjusted P=.035) and differed in racial composition (adjusted P<.001).

Among participants invited to follow-up, none of the examined baseline demographic or symptom comparisons between respondents (n=423) and nonrespondents (n=84) survived false discovery rate correction (all adjusted P>.25). Availability of at least one follow-up assessment differed by condition within this invited cohort (χ²₃=18.76, P<.001): 87.5% for guided journaling, 89.8% for unguided journaling, 86.7% for mood check only, and 72.5% for passive control.

#### Primary arm results

Baseline-adjusted mixed-effects ANCOVA models examined each intervention arm relative to passive control. Directional one-tailed tests were used for the guided and unguided journaling arms, whereas the mood tracking-only arm was evaluated using two-tailed tests. Models included baseline symptom scores as covariates and participant-level random intercepts to account for repeated observations and incomplete follow-up data under restricted maximum likelihood estimation.

At week 12 (1-month follow-up), guided journaling (Group 1) had the largest estimated anxiety difference favoring intervention relative to control (β = −1.78, d = 0.25, one-tailed p = 0.030), followed by unguided journaling (Group 2; β = −1.22, d = 0.17, one-tailed p = 0.097), while the mood tracking–only condition (Group 3) showed minimal difference from control (β = −0.48, two-tailed p = 0.615). The corresponding two-tailed P values for guided and unguided journaling were .060 and .194 at week 12 (.233 and .350 at week 8); thus, none of the journaling contrasts met the conventional two-sided α=.05 threshold. No arm-level contrasts reached statistical significance at week 8. No intervention effects were observed for depression or impairment (i.e., WHODAS) across arms or timepoints. To account for multiplicity, false discovery rate (FDR) correction was applied separately within each outcome across the six arm-level contrasts (three intervention arms × two follow-up assessments). No individual arm comparison met the α = 0.05 threshold after correction; for guided journaling at week 12, pFDR = 0.179.

Under the specified priors, Bayesian ANCOVA analyses estimated a modest reduction in anxiety in the journaling arms, strongest for guided journaling at week 12 (posterior mean = −1.79, 90% HDI [−3.33, −0.23], probability of decrease ≈97%). Evidence for unguided journaling was weaker (posterior mean = −1.22, 90% HDI [−2.77, 0.35], probability ≈90%), and mood tracking showed no benefit. Posterior estimates for depression were centered near zero across arms and timepoints. Overall, arm-level results showed a consistent directional pattern favoring journaling for anxiety, although individual contrasts did not survive multiplicity correction in the frequentist analyses.

#### Pooled journaling analysi

A secondary analysis compared the combined journaling conditions with the combined non-journaling conditions (Figure 3). Guided-versus-unguided differences were not statistically significant (week-12 anxiety: β = −0.65, p = 0.465; depression: p = 0.44); this does not establish equivalence between the journaling approaches.

**Figure 3.**
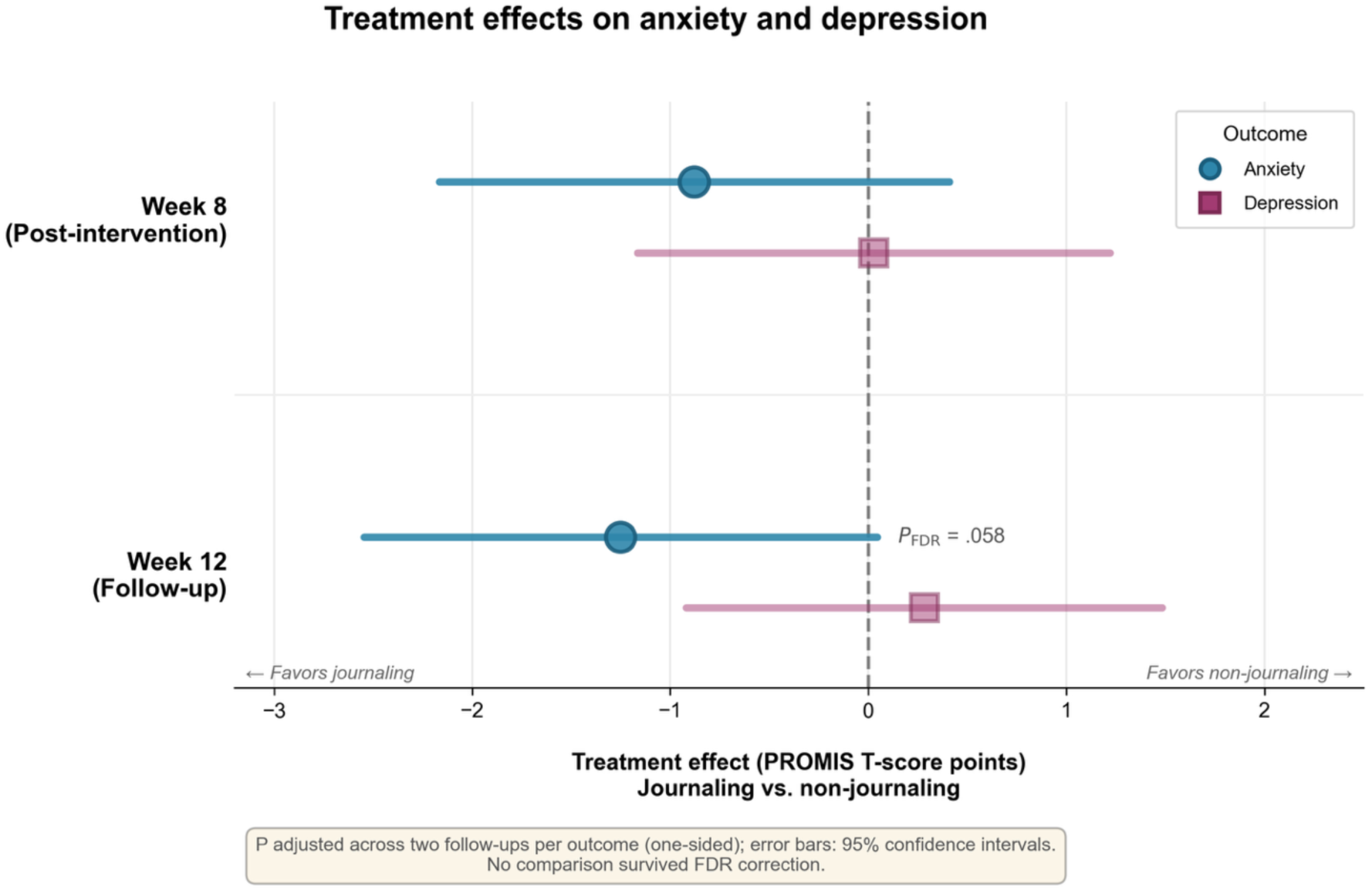
**Clinical** outcome estimates for **journaling on anxiety and depression.** Baseline-adjusted treatment-effect estimates comparing journaling (guided + unguided) with non-journaling conditions at post-intervention (week 8) and one-month follow-up (week 12). Points represent estimated between-group differences in PROMIS T-scores from linear mixed-effects ANCOVA models; error bars indicate 95% confidence intervals (414 participants with at least one PROMIS follow-up from the 498-person contamination-corrected cohort). Anxiety estimates favored journaling, but no contrast survived correction for multiple comparisons; no detectable effect was observed for depression.

Baseline-adjusted mixed-effects models estimated lower anxiety among participants assigned to journaling conditions at both follow-ups. The week-8 effect was modest (β = −0.88, SE = 0.66, one-tailed p = 0.091), while the week-12 effect was larger (β = −1.25, SE = 0.66, one-tailed p = 0.029) but did not remain significant after FDR correction across the two follow-up comparisons (pFDR = 0.058). Estimated effect sizes were small (week 8 d ≈ 0.16; week 12 d ≈ 0.19). No evidence of benefit was observed for depression.

Bayesian analyses of the pooled contrast were directionally consistent, estimating a modest reduction in anxiety at week 12 (posterior mean = −1.26, 90% HDI [−2.29, −0.12], probability of decrease ≈ 97%) and weaker evidence at week 8 (posterior mean = −0.89, 90% HDI [−1.96, 0.19], probability ≈ 91%). Posterior estimates for depression remained centered near zero.

Together, the analyses characterized uncertain anxiety differences in the hypothesized direction, largest at week 12. No frequentist contrast survived multiplicity correction, and the corresponding Bayesian posteriors placed 90%–97% probability on effects in the hypothesized direction under the specified priors. Because pd and the one-sided P value are closely related quantities, these are two expressions of the same estimate rather than independent lines of evidence.

#### Sensitivity and robustness analyses

Intention-to-treat sensitivity analyses used all 665 assigned participants for cohort accounting and preserved original condition; 423 participants supplied 766 follow-up observations per PROMIS outcome. Reconciliation of the assignment roster supplied no additional follow-up observations, so adding these records left the previously fitted ITT estimates, standard errors, confidence intervals, and P values unchanged. Baseline PROMIS T-score conversions were checked against all 507 previously scored records, with exact agreement. In the four-arm ITT model, no arm-level contrast survived correction for multiple comparisons. At week 12, guided journaling had the largest estimated difference favoring intervention in anxiety relative to passive control (β = −1.48, SE = 0.94, one-tailed P = .057; corresponding two-tailed P = .114; pFDR = .343), followed by unguided journaling (β = −0.93, SE = 0.93, one-tailed P = .158; pFDR = .473), while mood tracking showed minimal difference from control (β = −0.20, SE = 0.94, two-tailed P = .833; pFDR = .833). No evidence of benefit for depression was observed in the arm-level comparisons.

In the secondary pooled ITT analysis, journaling versus non-journaling conditions remained directionally favorable for anxiety, although neither follow-up comparison survived FDR correction (week 8: β = −0.62, SE = 0.66, one-tailed P = .174, pFDR = .174; week 12: β = −1.10, SE = 0.66, one-tailed P = .048, pFDR = .095). Corresponding two-tailed P values were .348 and .095, respectively. The week-12 pooled anxiety estimate was −1.10 T-score points (95% CI −2.40 to 0.19). Additional adjustment of the ITT models for years of education, race and ethnicity, or both left treatment estimates materially unchanged (largest absolute coefficient change, 0.18 T-score points), and no contrast changed significance status after false discovery rate correction. These available-data comparisons did not establish efficacy.

Additional post hoc contrasts from the four-arm ITT model compared each journaling condition directly with mood tracking alone. Anxiety estimates favored both guided and unguided journaling at both assessments. At week 12, estimated differences were −1.28 PROMIS T-score points for guided journaling (95% CI −3.13 to 0.57) and −0.74 for unguided journaling (95% CI −2.57 to 1.10). All confidence intervals included zero, and no comparison survived FDR correction across six contrasts per outcome: guided journaling, unguided journaling, and passive control versus mood tracking at two follow-ups. Depression estimates did not favor journaling.

In the pooled active-comparator sensitivity analysis, we compared the pooled journaling conditions directly with the mood-check-only condition, excluding passive control. Anxiety estimates remained in the hypothesized direction but were smaller and did not reach statistical significance (week 8: β = −0.78, SE = 0.79, one-tailed P = .162, corresponding two-tailed P = .325, pFDR = .162; week 12: β = −1.04, SE = 0.80, one-tailed P = .097, corresponding two-tailed P = .194, pFDR = .162). No benefit was observed for depression. Thus, the direction of the anxiety signal was preserved when journaling was compared with another active app condition rather than passive control, although the comparison remained statistically uncertain.

As an additional model-specification sensitivity analysis, conventional complete-case ANCOVA produced anxiety estimates in the same direction (week 8: β = −1.02, SE = 0.65, one-tailed P = .059, pFDR = .089, d = 0.145; week 12: β = −0.91, SE = 0.68, one-tailed P = .089, pFDR = .089, d = 0.131). No corresponding benefit was observed for depression. These alternative specifications also left the anxiety estimates statistically uncertain; they do not address bias from unobserved follow-up outcomes.

Finally, in a post hoc analysis, we assessed the sensitivity of the Bayesian estimates to the prior specification (Supplementary Table S4, Supplementary Figures S3 and S4). The direction of the anxiety estimate was stable across all priors examined: every journaling contrast remained negative at both follow-ups under every specification. At week 12, the posterior probability of symptom reduction was ≥0.95 for the pooled contrast, ≥0.92 for guided journaling, and ≥0.80 for unguided journaling, which was the least stable. The magnitude was prior-dependent: the week-12 guided-journaling estimate ranged from −1.79 under a near-flat prior to −0.94 under the most skeptical specification. Under that skeptical prior all three journaling estimates fell within the range used for practical equivalence in this post hoc analysis (±1 T-score point; guided −0.94, unguided −0.54, pooled −0.91), with 53–75% of posterior mass inside it.

#### Engagement patterns and distress-dependent response

Exploratory analyses indicated that overall engagement metrics (entry frequency, compliance, word count) did not predict 12-week anxiety outcomes after baseline adjustment (all p > 0.38). Average immediate post-session mood improvement showed no significant main effect in the full sample (β = 0.85, p = 0.12).

However, distress level, as indexed by automated risk classification, significantly moderated the association between immediate mood improvement and long-term anxiety reduction (immediate improvement × distress: β = 2.54, p = 0.018). Stratified analyses indicated that immediate improvement predicted 12-week anxiety reduction among users with concerning content (β = 2.08, p = 0.012, controlling for baseline; N = 71), but not among non-distressed (p = 0.88; N = 30) or high-risk users (p = 0.34; N = 14).

At the session level, word count was modestly associated with immediate mood improvement (β = 0.082, p = 0.018) but did not predict long-term anxiety outcomes in any distress stratum (all p > 0.26). Non-distressed users exhibited larger mean immediate mood improvements than distressed users (F(2,152) = 4.29, p = 0.016), yet immediate response was only predictive among the distressed group. Within the unguided arm, participants with concerning or high-risk entries demonstrated higher retention than those without elevated risk flags (X^2^(2) = 8.14, p = 0.017).

These exploratory subgroup associations suggest questions for prospective evaluation; they do not establish that immediate mood change predicts treatment benefit or validates selection of particular users for journaling.

#### PART II. Measurement of Mood and Clinically Relevant Risk Dynamics

The nine participants with documented crossover had been randomized to passive control and therefore did not contribute to Part II behavioral analyses, which were restricted to active-arm data. Trial-based text-derived risk analyses were restricted to the unguided journaling arm because automated risk classification was consistently available for entries in that condition; risk-classification availability varied across the prompts used in guided journaling.

#### Age-Level risk prevalence

High-risk entries were modestly more common among younger users (Figure 4). A grouped-binomial logistic regression model with robust (HC3) standard errors showed a significant negative association between age and the probability that a journal entry was classified as high-risk (β = −0.22, 95% CI [−0.39, −0.05], p = .013), corresponding to an odds ratio of 0.81 per year of age. This pattern indicates a gradual decline in high-risk entry likelihood across the late-adolescent to young-adult age range. Empirical age-specific means were broadly consistent with the modeled trajectory, although variability increased at older ages due to smaller sample sizes. No association was observed between high-risk entry frequency and sex, and age was not significantly associated with the likelihood of concerning entries. A comparable age-related gradient in high-risk entry rates was observed in the independent general-population dataset, supporting generalizability of the association.

**Figure 4.**
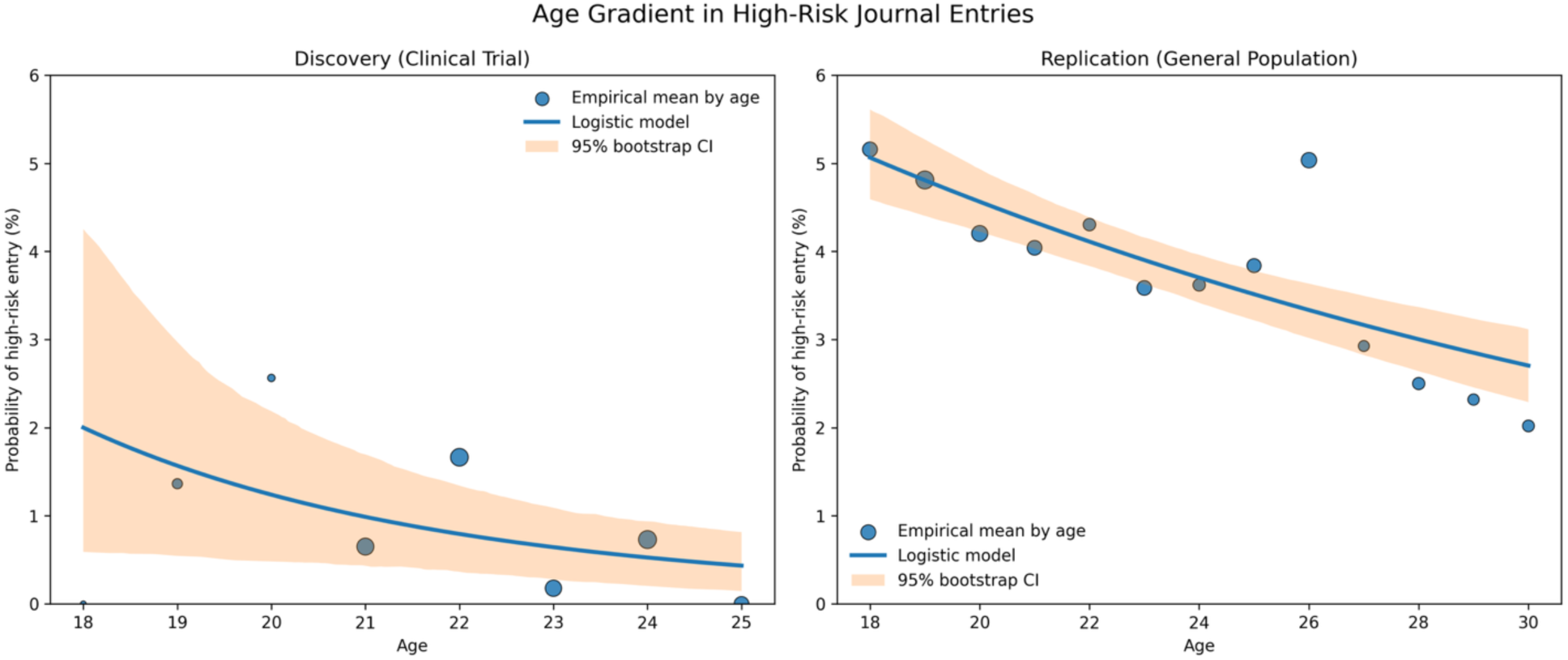
Age gradient in high-risk journal entries. Population-averaged probability that a journal entry is classified as high risk as a function of user age. Left: discovery dataset from the clinical trial. Right: replication in a general-population sample. Lines represent logistic model predictions; shaded bands indicate 95% bootstrap confidence intervals. High-risk classification probability decreases with increasing age.

#### Temporal patterns in affective and risk levels

Temporal analyses indicated clear time-of-day structure in affective states and distress risk signals (Figure 5). Hour-of-day models demonstrated significant time-of-day variation in distress risk prevalence (concerning or high-risk entries; population-averaged GEE p = 0.036) and strong time-of-day structure for energy (GEE p < 1x10^-12^), whereas pleasantness showed no significant time-of-day variation (GEE p = 0.22).

**Figure 5.**
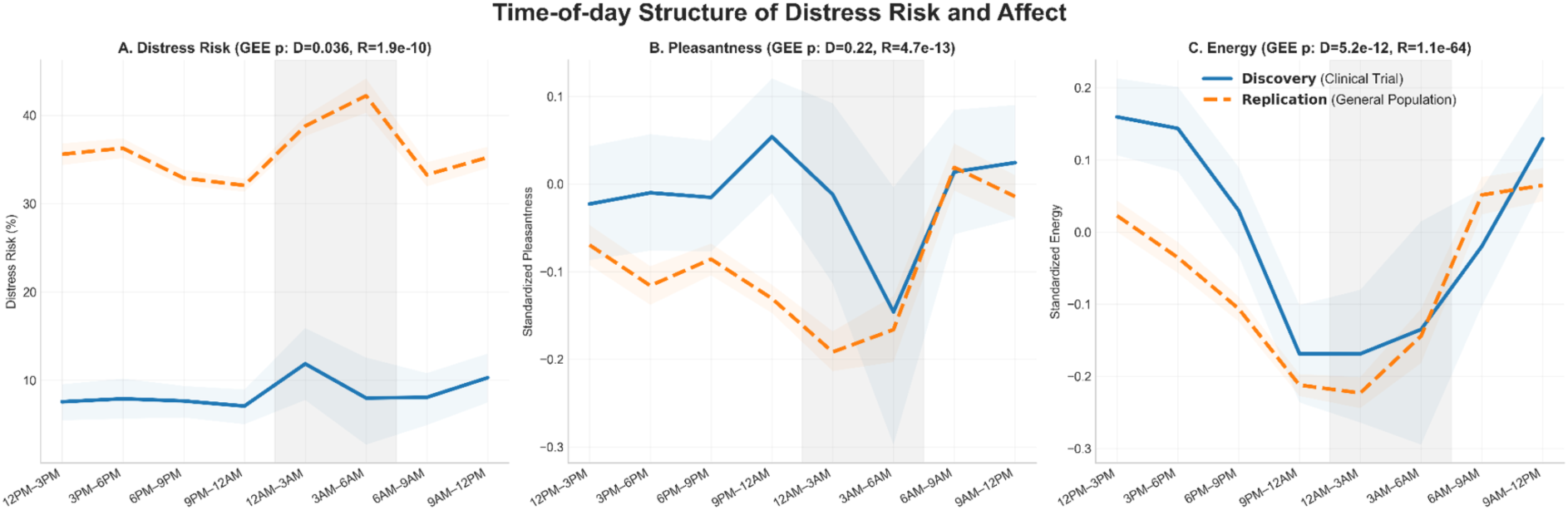
Time-of-day structure for distress risk and affect. Population-averaged temporal profiles across eight 3-hour time bins. Solid lines represent the clinical trial dataset (discovery) and dashed lines represent the general-population dataset (replication). Distress risk probability and energy exhibit significant time-of-day variation, whereas pleasantness shows minimal time-of-day structure. Shaded regions represent 95% clustered bootstrap confidence intervals. GEE models with exchangeable correlation structure were used for statistical inference. Overnight and early-morning estimates should be interpreted cautiously because these bins were relatively sparsely sampled.

Risk probability was highest during late-night and overnight hours (≈12% at 00:00–03:00), declined through early morning (≈8% at 03:00–06:00), showed a modest midday elevation (≈10% at 09:00–12:00), and remained lower during afternoon and evening periods (≈7–8%). Energy demonstrated a pronounced time-of-day pattern, rising from overnight lows to a midday peak before declining again in the evening. Pleasantness levels varied little across the day and did not show statistically reliable time-of-day structure.

These time-of-day patterns were broadly consistent in the independent, general-population dataset (Figure 5). In particular, the energy rhythm showed strong replication across datasets, with lowest levels occurring overnight and recovery through morning hours. Risk prevalence also showed similar overnight elevation in the replication sample, although absolute rates were higher in the general-population dataset. Because overnight and early-morning bins contained relatively few observations, absolute estimates during these periods should be interpreted cautiously.

Day-of-week analyses showed weaker and less consistent structure. In the clinical trial dataset, pleasantness exhibited a weekly rhythm (GEE p = 9.05×10⁻⁷) and energy showed a modest weekly effect (GEE p = 0.015), whereas risk flags did not cluster by day of week (GEE p = 0.24). However, these weekly patterns did not replicate in the general-population dataset, suggesting that day-of-week effects were less stable across contexts. However, previous work in structured diary ratings have shown interactions between day of week and time of day that were not examined here due to sample size.

Together, these findings indicate that acute distress signals are more closely aligned with time-of-day fluctuations in arousal and energy than with weekly social rhythms.

#### Transition probability and risk persistence

After exclusion of entries lacking valid affect ratings, the mood dataset contained 12,553 user × day × hour observations contributed by 357 active-arm users. Mood-transition pairs were constructed by linking each observation to the immediately subsequent observation from the same user occurring within ≤48 hours, yielding 10,227 eligible transitions from 317 users, with a median inter-entry interval of 24 hours. Because automated risk classification was available only in the unguided journaling condition, risk-state transitions were analyzed separately within that arm and restricted to observations with evidence that automated classification had occurred. This yielded 2,640 eligible ≤48-hour risk transitions from 99 users, with a median inter-entry interval of 24 hours. Figure 6 displays the corresponding entry-weighted transition matrices, while persistence estimates were calculated at the user level to avoid overweighting highly active participants. Transition matrices demonstrated significant non-random structure for both mood quadrants (χ²(9) = 1189.9, p < 10^-15^) and clinical risk states (χ²(4) = 110.2, p < 10^-15^), indicating that next-entry states were strongly conditioned on prior states rather than independent of prior state. These tests were conducted on transition counts and do not account for within-user dependence, and should therefore be interpreted descriptively.

**Figure 6.**
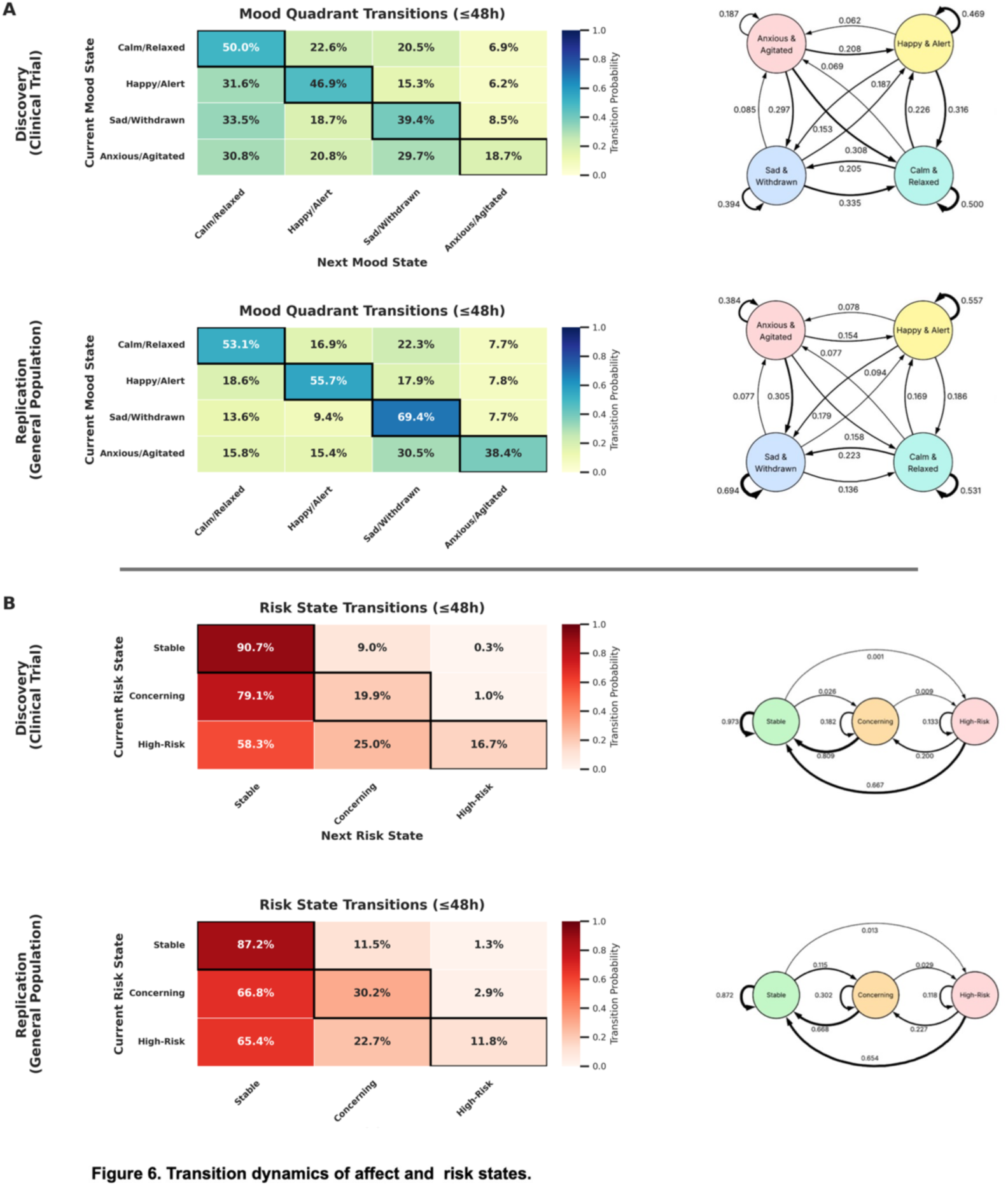
Transition dynamics of affect and risk states. Transition probability matrices showing conditional probabilities of the next entry state given the current state. Discovery (clinical trial) and replication (general population) datasets are shown. 6a. Mood quadrants demonstrate graded persistence with transitions favoring adjacent states rather than polarity reversals. 6b. Risk states show high stability for stable entries and low recurrence of high-risk states.

To avoid over-weighting highly active users, persistence estimates were computed using user-cluster bootstrap resampling (1,000 iterations), with each participant contributing equally. Mood states exhibited graded persistence across successive entries. Calm/Relaxed showed the highest persistence (41.5%, 95% CI [38.7%, 44.3%]), followed by Happy/Alert (31.7%, 95% CI [28.4%, 35.0%]) and Sad/Withdrawn (26.8%, 95% CI [23.9%, 29.9%]), whereas Anxious/Agitatedexhibited the lowest persistence (13.2%, 95% CI [10.2%, 16.3%]), consistent with greater short-term instability in high-arousal negative affect.

Clinical risk states also showed clear persistence structure. Stable entries remained Stable at the subsequent eligible entry in 88.9% of transitions (95% CI [85.7%, 91.7%]). Concerning states exhibited substantially lower persistence (12.4%, 95% CI [8.5%, 16.7%]), most frequently transitioning back to Stable states. High-Risk states showed modest persistence (9.4%, 95% CI [0.0%, 21.9%]), although estimates were imprecise because only 8 users contributed eligible transitions from a High-Risk state. Accordingly, High-Risk persistence estimates should be interpreted descriptively. Automated risk classifications were available only in the unguided journaling arm.

Across both affective and clinical domains, next-entry states exhibited clear short-term state dependence, consistent with structured transition dynamics. A similar transition structure was observed in the independent general-population dataset (Figure 6), supporting the reproducibility of these transition dynamics across the two datasets.

### Volatility and acute drop susceptibility

#### Drop definition and volatility structure

To determine whether affective instability predicts abrupt mood shifts, we examined whether volatility in pleasantness and energy was associated with susceptibility to acute affective drop events. Acute drop events were defined as ≥3-point decreases (30% of the mood scale range) in daily mean scores within a window of four successive observed dates; sensitivity analyses examined thresholds from ≥2 to ≥5 points. Among users with ≥10 observations (n = 289), both valence and energy drops were common but showed substantial inter-individual variability.

Energy and pleasantness volatility (residualized for mean levels) were moderately correlated (r = 0.59, p < 10⁻²⁷; Figure 7A), indicating partially overlapping but distinct instability phenotypes. Multivariable models therefore examined dimension-specific predictive effects.

**Figure 7.**
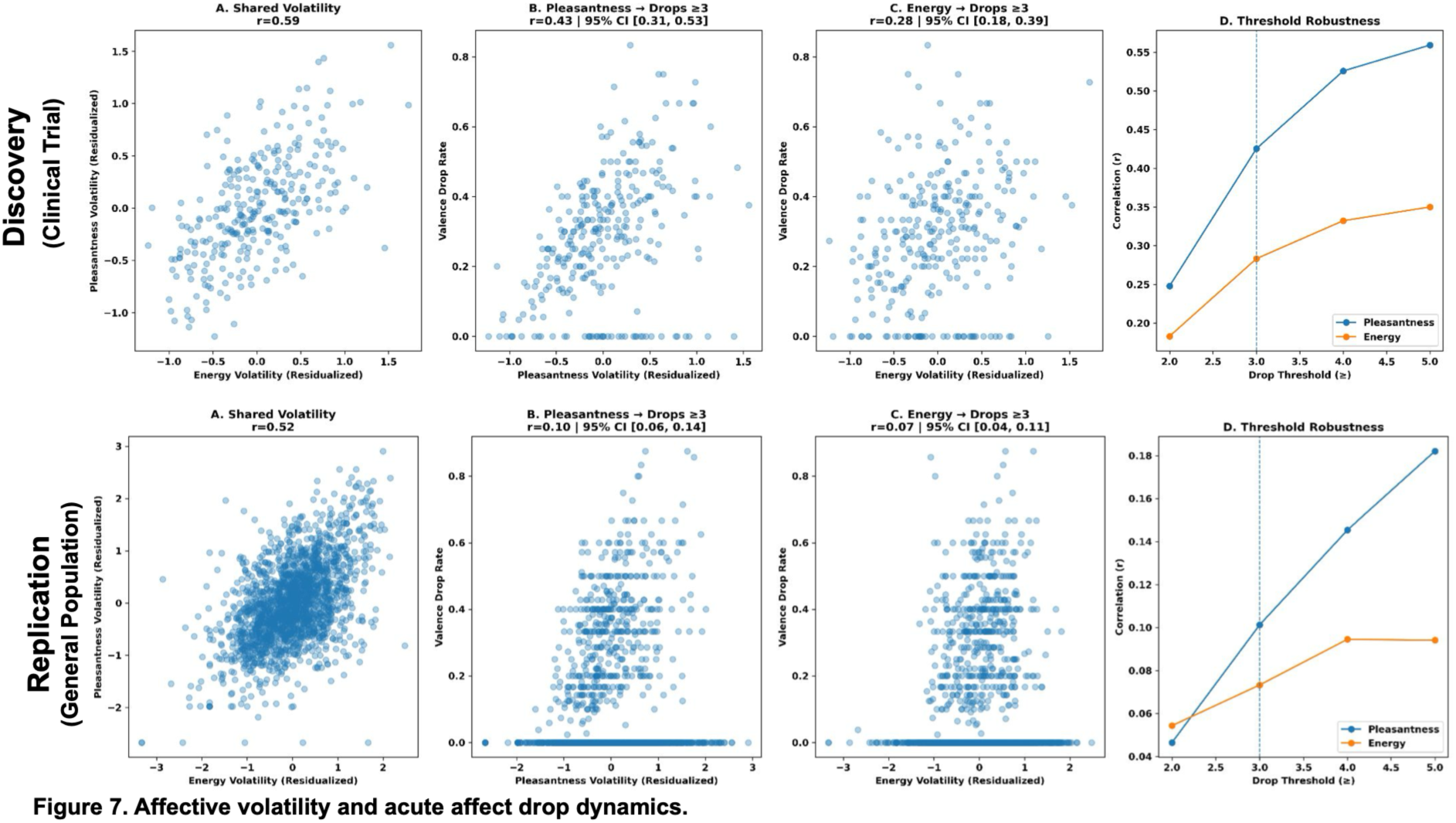
Affective volatility and acute affect drop dynamics. Associations between user-level volatility and susceptibility to acute affective drops. Volatility measures were computed as residualized within-user standard deviations for pleasantness and energy. Discovery (clinical trial) and replication (general population) datasets are shown. Pleasantness and energy volatility share variance; panels B–D show their associations with valence-drop rate. Correlations increase with larger drop thresholds.

#### Dimension-specific associations

Pleasantness volatility was associated with valence-drop frequency at the ≥3-point threshold (r = .43, 95% CI [.31, .54], P < .001), whereas energy volatility showed a weaker cross-axis association (r = .28, 95% CI [.17, .39], P < .001). In sensitivity analyses restricted to users with sufficient data to estimate both volatility and drop rate (n=253), these associations strengthened to r=.65 and r=.39, respectively. However, temporal permutation analyses of this complete-pair sample produced correlations of similar magnitude under both unrestricted and block-preserving shuffles. Thus, the association between volatility and threshold-defined drop frequency primarily reflects the expected relationship between overall affective variability and susceptibility to large excursions, rather than additional information carried by the temporal ordering of observations.

When drops were defined on the energy axis, effects were again dimension-specific. Energy volatility was associated with energy-drop frequency (r = 0.36, 95% CI [0.25, 0.46]), whereas pleasantness volatility showed weaker cross-axis association. In multivariable models at the ≥3-point threshold, energy volatility remained the only significant volatility predictor.

#### Threshold robustness and cross-axis coupling

In the analyses shown in Figure 7D, correlations increased monotonically with drop threshold. Correlations between pleasantness volatility and valence-drop rate rose from r = 0.25 (≥2 points) to r = 0.43 (≥3), r = 0.53 (≥4), and r = 0.56 (≥5). Correlations between energy volatility and valence-drop rate increased from r = 0.18 (≥2) to r = 0.28 (≥3), r = 0.33 (≥4), and r = 0.35 (≥5), indicating stronger relationships at larger drop thresholds.

Event-level coupling was modest: valence-drop depth was weakly associated with energy decreases within the same window (r = 0.15), and energy-drop depth with pleasantness decreases within the same window (r = 0.12).

Volatility metrics were not associated with subsequent high-risk classifications in logistic regression models accounting for clustering within users. Overall, affective instability was associated with susceptibility to acute within-axis drops. Associations of both volatility measures with valence-drop susceptibility were observed in the independent, general-population dataset, although correlations were smaller (Figure 7).

##### Risk escalation patterns

To characterize short-term escalation dynamics, we estimated the probability that the next journal entry would be classified as high-risk or concerning conditional on prior distress history (Figure 8), using sequential entry pairs within users. Six history definitions were evaluated: ≥1 high-risk entry within the prior three entries, ≥1 high-risk entry within the prior 72 hours, any prior high-risk entry during the observation period, and parallel definitions for concerning entries. Outcome probabilities were estimated using a user-level rate estimator (equal weighting across users) with bootstrap resampling over users (2,000 iterations) to generate 95% confidence intervals.

**Figure 8.**
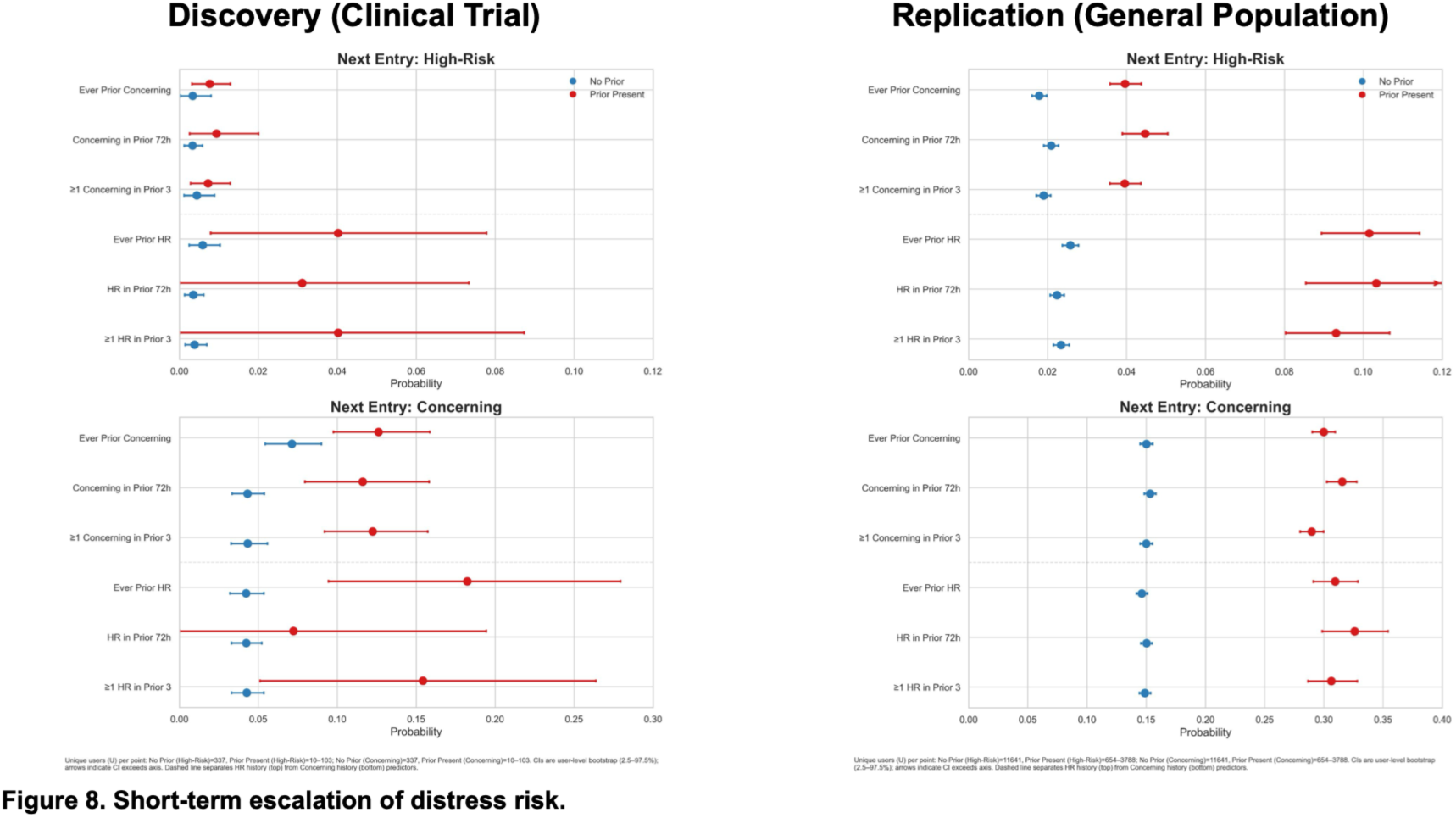
Short-term escalation of distress risk. User-weighted estimates of the probability that the next entry is classified as high risk or concerning conditional on prior high-risk history. Discovery (clinical trial) and replication (general population) datasets are shown. Prior high-risk entries are associated with increased probability of subsequent elevated-risk states, indicating short-term persistence and graded escalation of distress signals.

Prior distress history was associated with elevated probabilities of both subsequent high-risk and concerning entries, although estimates for high-risk predictors were less precise due to the relatively small number of high-risk observations. High-risk history showed the strongest effects: when no prior high-risk history was present, the probability of the next entry being classified as high-risk was ≈0.4–0.7%, increasing to ≈3–4% when prior high-risk events were present. Concerning history produced smaller increases in high-risk probability (≈0.4% to ≈1.2–1.5%).

For concerning outcomes, probabilities increased from ≈4–5% without prior high-risk history to ≈7–18% when prior high-risk events were present, with concerning history producing intermediate elevations (≈5% to ≈10–12%).

These results indicate short-term persistence and graded escalation across distress categories, with escalation more often manifesting as concerning entries rather than immediate recurrence of high-risk classifications. This pattern suggests that acute high-risk events typically transition into sustained elevated distress rather than repeating as immediate high-risk states.

Consistent with this interpretation, analyses of affective antecedents showed that high-risk entries were preceded by lower mean pleasantness and lower mean energy, whereas short-term volatility differences were modest and not significant in multivariable models. Together, these findings suggest that high-risk states arise primarily from sustained low affect and persistent distress rather than isolated volatility spikes. Similar escalation patterns were observed in the independent, general-population dataset (Figure 8), supporting the generalizability of these dynamics across contexts.

## DISCUSSION

This study evaluated a privacy-preserving digital journaling platform as both a low-intensity mental health intervention and a scalable behavioral measurement system. In Part I, the randomized trial did not establish a statistically significant clinical benefit of guided or unguided journaling after correction for multiple comparisons. Anxiety estimates were small and consistently in the hypothesized direction, with the largest effects observed at the week-12 follow-up, but no arm-level or pooled contrast survived multiplicity correction. We found no evidence of benefit for depression or functional impairment. Part II addressed a separate, exploratory, non-preregistered objective and identified reproducible time-of-day, persistence, and escalation patterns across the trial and an independent general-population dataset. These findings do not alter the interpretation of the randomized clinical outcomes; they speak instead to the potential of privacy-preserving journaling data for longitudinal behavioral measurement and classifier-derived risk-signal characterization.

Digital journaling should be considered within a rapidly expanding ecosystem of low-intensity, technology-enabled mental health supports. This ecosystem includes psychoeducational video curricula, self-guided wellness applications, single-session digital interventions, and conversational agents[42–44]. Journaling occupies a comparatively simple but potentially useful niche. It requires minimal instruction, aligns with naturally occurring patterns of occasional self-reflection, and integrates into mobile and web-based environments with flexible schedules[17]. Increasingly, journaling features are embedded within broader multifeature platforms. Guided journaling, including initial prompts or contextual nudges, may extend accessibility for users who are less comfortable with unstructured reflection[45]. In the present study, however, guided participants selected from a broad menu of general reflective prompts rather than a symptom-specific therapeutic protocol. Future trials should test whether prompts matched to particular symptoms or mechanisms, such as worry, avoidance, cognitive reappraisal, gratitude, positive affect, rumination, or behavioral activation, produce different outcomes. The present study cannot determine whether greater specificity would improve outcomes. The absence of demonstrated benefit for depression also motivates testing whether prompts addressing depression-related processes differ from general reflective writing; this trial did not test that comparison.[45]

From a service-delivery perspective, digital journaling may ultimately prove more useful as an accessible, low-intensity support than as a standalone treatment. The current findings, however, do not establish that role. Overall engagement intensity, including entry frequency, compliance, and word count, did not predict long-term anxiety reduction. Immediate post-session mood improvement also showed no main effect in the full sample. Its association with week-12 anxiety reduction was observed only among users with concerning classifier-derived content and arose from an exploratory subgroup analysis. This pattern is potentially informative, but it should be treated as hypothesis-generating rather than evidence that journaling benefited distressed users. Prospective studies will be needed to determine whether immediate response can identify users who may benefit from particular forms of guided reflection and whether journaling can support self-awareness, help-seeking, maintenance after structured treatment, or stepped-care pathways [20,46,47].

Beyond symptom change, Part II examined whether repeated, privacy-preserving observations could reveal clinically relevant behavioral structure. Recent and sustained low pleasantness was associated with next-entry distress, whereas short-term volatility contributed little additional predictive information. Time-of-day analyses identified variation in classifier-derived distress and self-reported energy, and transition analyses showed short-term persistence and graded escalation across mood and risk states. Selected patterns were reproduced in the independent general-population dataset, although absolute rates differed. Together, these findings suggest that short sequences of prior observations may contain information that is obscured by episodic symptom assessments [20,21]. Importantly, the analyses characterize temporal and population-level patterns in self-reports and automated proxy signals. They do not demonstrate prediction of clinical events, establish a clinical monitoring system, or support use of classifier outputs for clinical decision-making.

The platform’s privacy-preserving architecture is relevant to this measurement contribution. The reported signals were derived from structured mood ratings, classifier outputs, timestamps, word counts, and engagement metadata rather than retained journal text. This approach reduces the amount of sensitive content retained for research while allowing longitudinal patterns to be characterized. Privacy preservation, however, does not resolve questions of measurement validity. Although the classifier was evaluated against held-out clinician-rated entries, it was not directly validated against clinician-adjudicated entries from the trial or general-population samples. Because raw text was deleted after processing, retrospective in-domain validation was not possible. Classifier-derived categories should therefore be interpreted as operational proxies for distress and safety-relevant language rather than clinical diagnoses or ground truth. Population-specific calibration, subgroup evaluation, and prospective validation would be required before any clinical deployment[30,48,49].

Online panels offer a practical route to evaluating digital interventions outside clinical settings; recruitment through Prolific Academic yielded 665 assignments in approximately ten weeks. Participation in the app conditions, however, required moving from the panel environment into a separate application. Accepting a study and completing online assessments did not necessarily translate into installing, registering, and linking the app—a participation hurdle absent from studies conducted entirely within a survey platform. Of 184 participants assigned to app conditions who provided neither follow-up outcome, 141 (76.6%) had not completed onboarding or linkage and were therefore not invited to follow-up assessments. In passive control, 17 participants without follow-up were not invited because of administrative or enrollment-processing issues; the remaining 41 were invited but provided neither assessment. Despite similar week-12 follow-up availability across conditions (54.4%–58.7%), participation pathways differed substantially. Documented difficulties with Research IDs, account access, and assignment completion indicate procedural friction, although the design cannot distinguish its contribution from reluctance to install an external application or declining interest.

These differences underscore the value of an active app comparator. Comparisons of journaling with mood tracking alone, individually or pooled, shared onboarding requirements and yielded anxiety estimates in the hypothesized direction, although they remained statistically uncertain. These analyses assess the pattern under more comparable participation demands, while potential selection associated with engagement and follow-up completion remains relevant.

Future trials could use common neutral onboarding procedures across conditions, including controls, to establish comparable technical readiness before condition-specific activities begin. Participant flow should be anchored at assignment, with installation, registration, linkage, and first use recorded separately, and follow-up assessments accessible independently of app engagement. Refining this transition alongside more targeted journaling content would strengthen the foundation for larger, prospectively powered trials while preserving the recruitment efficiency of online panels.

Several limitations warrant consideration. First, the anxiety estimates were small and did not survive correction for multiple comparisons. Complementary Bayesian models characterized effect direction, magnitude, and uncertainty but should not be interpreted as independent confirmation of the frequentist results. Under more skeptical priors, estimated magnitudes attenuated toward the region of practical equivalence used in the post hoc sensitivity analysis, leaving the clinical relevance of the anxiety signal uncertain. For depression, posterior estimates remained concentrated near zero across prior specifications. Second, compensation, assigned activity, study contact, and engagement differed across conditions. All participants could receive up to $40 for assessment completion; additional activity-based compensation was up to $84 in the journaling conditions, up to $28 in the mood-check-only condition, and $0 in passive control. Participants, study personnel who sent reminders and provided technical support, and analysts were unmasked. These differences may have affected expectancy, participation, engagement, or retention, and the design cannot separate the effects of compensation, attention, assigned activity, and journaling content. Similar follow-up availability across conditions does not rule out selection bias in the available-data estimates if missingness depended on unobserved outcomes. Documented crossover was addressed in sensitivity analyses, but journaling outside the study platform and use of other mental health applications were not systematically assessed.

Additional limitations apply to Part II. Journal-entry times were unevenly distributed, and engagement intensity varied substantially. The behavioral analyses necessarily characterize participants who generated sufficient data and may not generalize to users with little or no platform engagement. High-risk classifications and next-entry high-risk events were uncommon, limiting precision. The association between volatility and threshold-defined affective drops was largely attributable to marginal affective variability rather than added information from temporal ordering. Finally, the trial sample consisted of young adults with mild-to-moderate symptoms, was not representative of the general population, and was not systematically assessed for histories of mental or behavioral disorders. Replication of selected behavioral patterns in the independent general-population dataset increases confidence that those patterns were not unique to the trial context, but it does not remove these limitations or establish clinical validity.

In conclusion, the randomized trial did not establish a clinical benefit of guided or unguided digital journaling for anxiety, depression, or functional impairment. The uncertain anxiety estimates and absence of demonstrated benefit for depression can inform subsequent studies building on prior expressive-writing research, including direct tests of symptom-focused guided-journaling approaches. Separately, Part II showed that privacy-preserving longitudinal journaling data could be used to characterize reproducible temporal patterns in mood and classifier-derived distress and risk signals. These findings support further evaluation of digital journaling as a behavioral measurement platform. The trial also showed that most participation loss in the app conditions occurred during onboarding or linkage, indicating that recruitment, onboarding, engagement, and outcome completion should be evaluated as distinct stages in digital trials. Future research should refine this transition, extend longitudinal follow-up, and prospectively validate classifier-derived signals before integrating these tools into clinician-monitored workflows.

## AUTHOR CONTRIBUTIONS

Conceptualization: MPM, JT, MK, LA, CW, AK, BL, MF, and GK;

Data curation: AE, SY, YW, LA, LH, and MSK;

Formal analysis: MPM, DML, AE, CJ, and GK;

Funding acquisition: MPM, HK, and BL;

Investigation: MK, SY, YW, LM, and RM;

Methodology: MPM, DML, GS, JT, MK, RVDW, GAS, KM, TDS, and GK;

Project administration: JT, MK, ML, LA, MSK, and MF;

Resources: MPM, GK, ARF, HK, and BL;

Software: MPM, DML, AE, and RVDW;

Supervision: MPM and GK;

Validation: MPM, JT, RVDW, BL, and GK;

Visualization: MPM, DML, AE, GK, and CJ;

Writing (original draft): MPM, DML, and GK;

Writing (review & editing): all authors. All authors critically reviewed the manuscript, approved the final version, and accept accountability for their respective contributions and for the integrity of the work.

## Data Availability

All data produced in the present study are available upon reasonable request to the authors

## ACKNOWLEDGEMENTS

Mirror Journal is part of a broader portfolio of scalable digital mental health tools developed by the Child Mind Institute. We thank AlphaSouth for software development of the Mirror platform, and ReD Associates for early-stage user research informing its design. We also acknowledge the use of large language models (ChatGPT, Claude, Gemini) for assistance with editing, drafting, and code development; all final content and analyses were reviewed and approved by the authors. MPM is the Phyllis Green and Randolph Cowen Scholar at the Child Mind Institute.

## Data Availability

Raw journal text was not retained and therefore cannot be shared. De-identified derived data and model outputs may be made available upon reasonable request, subject to institutional approval and applicable ethical and privacy constraints.

## Code Availability

Analysis code supporting the reported analyses is available from the corresponding author upon reasonable request.

## SUPPLEMENTARY MATERIALS

### SUPPLEMENTARY METHODS

#### Trial procedures and protocol modifications

The trial began with Mirror Journal version 1.0.4.

A Research ID feature was introduced between weeks 1 and 2 to link application activity to study records. An initial proof-of-entry procedure that requested image uploads was discontinued; the uploaded images were never viewed by investigators, were deleted, and were not used in any analysis. Entry completion was subsequently verified using application metadata. Operational issues included incorrect Research IDs, participants completing the wrong assignment, account recovery or replacement, and an assignment-submission error in approximately week 4; these were addressed through participant messages, troubleshooting, and account linkage using Research IDs. No privacy incidents were reported during the trial.

#### Study reminders and participant support

Participants were asked to enable application notifications, and Prolific Academic notified participants when weekly check-ins and post-study questionnaires became available. Study staff sent assignment reminders in week 1, targeted Research ID reminders in week 2, reminders to participants who had completed the wrong assignment in week 3, submission-error guidance in week 4, completion links to participants marked incomplete in week 5, and post-study compensation reminders.

#### Retrospective sensitivity analysis

No formal power analysis was specified in the trial registration; these calculations were performed after data collection and did not inform enrollment. Among the 665 assigned participants, 379 had paired baseline and week-12 PROMIS measurements (guided 95, unguided 98, mood check only 92, passive control 94). Arm-adjusted baseline–week-12 Pearson correlations were .596 for anxiety and .576 for depression. Using these complete-pair arm sizes, a baseline-adjusted ANCOVA approximation, 80% power, and an unadjusted one-sided α=.05, the approximate minimum detectable effect size was d=.29–.30 for guided or unguided journaling versus passive control and d=.21 for pooled journaling versus non-journaling. In the contamination-corrected subset, 372 complete pairs (95, 98, 92, and 87) and correlations of .606 for anxiety and .588 for depression produced the same rounded values. These calculations characterize week-12 complete-pair comparisons, not power for the observed effect or the full longitudinal mixed-effects models. They do not incorporate multiplicity correction or account for selection from missing follow-up.

#### Bayesian ANCOVA

To estimate intervention effects on PROMIS anxiety and depression T-scores, we fit Bayesian linear mixed-effects ANCOVA models separately for each outcome. Models adjusted for baseline symptom severity and included fixed effects for treatment group, timepoint (Week 8, Week 12), and their interaction, with a random intercept for participant:

outcome ~ baseline + timepoint × group + (1 | participant)

Control and Week 8 served as reference levels. Weakly informative priors were specified based on the PROMIS scale (intercept ∼ Normal(50, 20); baseline coefficient ∼ Normal(0.5, 0.3); other coefficients ∼ Normal(0, 10); residual SD ∼ HalfNormal(10); participant random intercept ∼ Normal(0, sd) with sd ∼ HalfNormal(s), s set automatically by bambi from the dispersion of the outcome and the scales of the common-effect priors). Models were estimated in Python (bambi, pymc) using the No-U-Turn Sampler (4 chains; 2,000 warmup and 2,000 posterior draws per chain), with convergence confirmed (R-hat ≤ 1.01; adequate effective sample sizes).

Posterior mean differences versus control were summarized using 90% highest density intervals (HDI), reported throughout for consistency with the directional framing of the intervention hypotheses. For the journaling contrasts, which were evaluated directionally, we report the posterior probability of symptom reduction, P(effect < 0). For the mood tracking-only contrast, which was evaluated non-directionally, we report the probability of direction, that is, the posterior probability mass on the larger side of zero. Both are reported as descriptive summaries rather than as independent tests, alongside the proportion of posterior mass within a region of practical equivalence (ROPE; ±1 T-score point). A secondary analysis fitted a separate model in which guided and free-form journaling were pooled into a single “journaling” condition and compared against non-journaling (mood tracking + control), with non-journaling as the reference condition.

## SUPPLEMENTARY RESULTS

### Component-Model Performance

The component models demonstrated different operating characteristics consistent with their intended roles. For high-risk classification, LlamaGuard showed greater sensitivity than the RoBERTa high-risk classifier but lower specificity. RoBERTa applied a more conservative high-risk threshold, with higher specificity and precision but lower sensitivity.

For concerning classifications, the prompted Llama model demonstrated higher sensitivity and F1 performance than the RoBERTa concerning classifier, with lower specificity and precision. This pattern was consistent with the prompted model’s intended role as the principal detector of broader distress and emerging concern, while RoBERTa provided an additional suicide- and self-harm–focused classification pathway.

**Table S1.** Component Model Performance.

| Model | Reference category | Sensitivity | Specificity | Precision | F1 |
|---|---|---|---|---|---|
| LlamaGuard | High risk | 0.920 | 0.388 | 0.689 | 0.788 |
| RoBERTa | High risk | 0.648 | 0.812 | 0.835 | 0.730 |
| Prompted Llama | Concerning | 0.918 | 0.688 | 0.973 | 0.944 |
| RoBERTa | Concerning | 0.469 | 0.938 | 0.989 | 0.636 |

***Note:*** *For LlamaGuard and RoBERTa high-risk classification, all false-positive cases were clinician-rated as concerning entries rather than no-concern entries.*

Qualitative review indicated that false-positive classifications followed identifiable patterns rather than appearing arbitrary: at the component level, LlamaGuard produced 52 false-positive high-risk classifications and RoBERTa produced 16; in every case, the entry was nevertheless clinician-rated as concerning rather than no concern. The prompted Llama model produced five false-positive concerning classifications. Two reflected broader distress or safety-relevant circumstances involving bereavement and concern about harm to a pet, whereas three involved contextual or non-distress material, including wordplay, political discussion, and humor. RoBERTa produced one false-positive concerning classification, involving concern about a pet rather than personal risk. Overall, the predominant false-positive pattern was overestimation of severity or difficulty resolving contextual attribution, rather than detection of clinically irrelevant content.

### Hierarchical Ensemble Performance

Applying the hierarchical ensemble to the 210-entry test set yielded an overall classification accuracy of 67.6%. For high-risk classification, the ensemble achieved sensitivity of 0.952, specificity of 0.365, precision of 0.688, and an F1 score of 0.799. Its comparatively low high-risk specificity primarily reflected escalation of clinician-rated concerning entries: all 54 false-positive high-risk classifications were clinician-rated as concerning rather than no concern.

The ensemble showed lower exact-category sensitivity for concerning entries (0.174), largely because 54 of the 69 clinician-rated concerning entries were elevated to the high-risk category under the safety-oriented hierarchical rule. When high-risk and concerning classifications were combined as detection of any clinically meaningful signal, the ensemble achieved sensitivity of 0.985, specificity of 0.688, precision of 0.974, and an F1 score of 0.980.

### Bayesian ANCOVA

#### Model Fit and Sample

Of the 507 participants in the original extraction, 498 remained after the contamination exclusion, with 414 contributing to both PROMIS outcome models. These models do not include all 665 assigned participants.

Bayesian analyses broadly mirrored the primary frequentist findings (Figure S1). All models converged satisfactorily (R-hat ≤ 1.01). Baseline symptom severity was a strong predictor of follow-up outcomes (anxiety β = 0.69; 90% HDI [0.61, 0.76]; depression β = 0.71; 90% HDI [0.63, 0.79]).

Guided journaling showed the strongest reductions in anxiety relative to control. At Week 8, the posterior mean difference was −1.12 (90% HDI: −2.65 to 0.37; pd = 0.89), indicating a directional but inconclusive signal. By Week 12, the estimate increased to −1.79 (90% HDI: −3.33 to −0.23; pd = 0.97). Free-form journaling showed a similar but weaker pattern at Week 12 (mean = −1.22; pd = 0.90). Mood tracking showed no benefit at either timepoint (pd ≤ 0.70; substantial posterior mass within the ROPE). In pooled analyses, journaling (guided + free-form) yielded anxiety estimates consistently in the hypothesized direction, with larger estimates at week 12 (mean = −1.26; 90% HDI excluding zero; pd = 0.97).

No intervention showed meaningful effects on depression. Posterior estimates were small (−0.36 to +0.31 T-score points), with high proportions of posterior mass within the ROPE (typically 70– 90%) and pd values ≤ 0.66, consistent with practical equivalence rather than inconclusive null effects.

**Figure S1.**
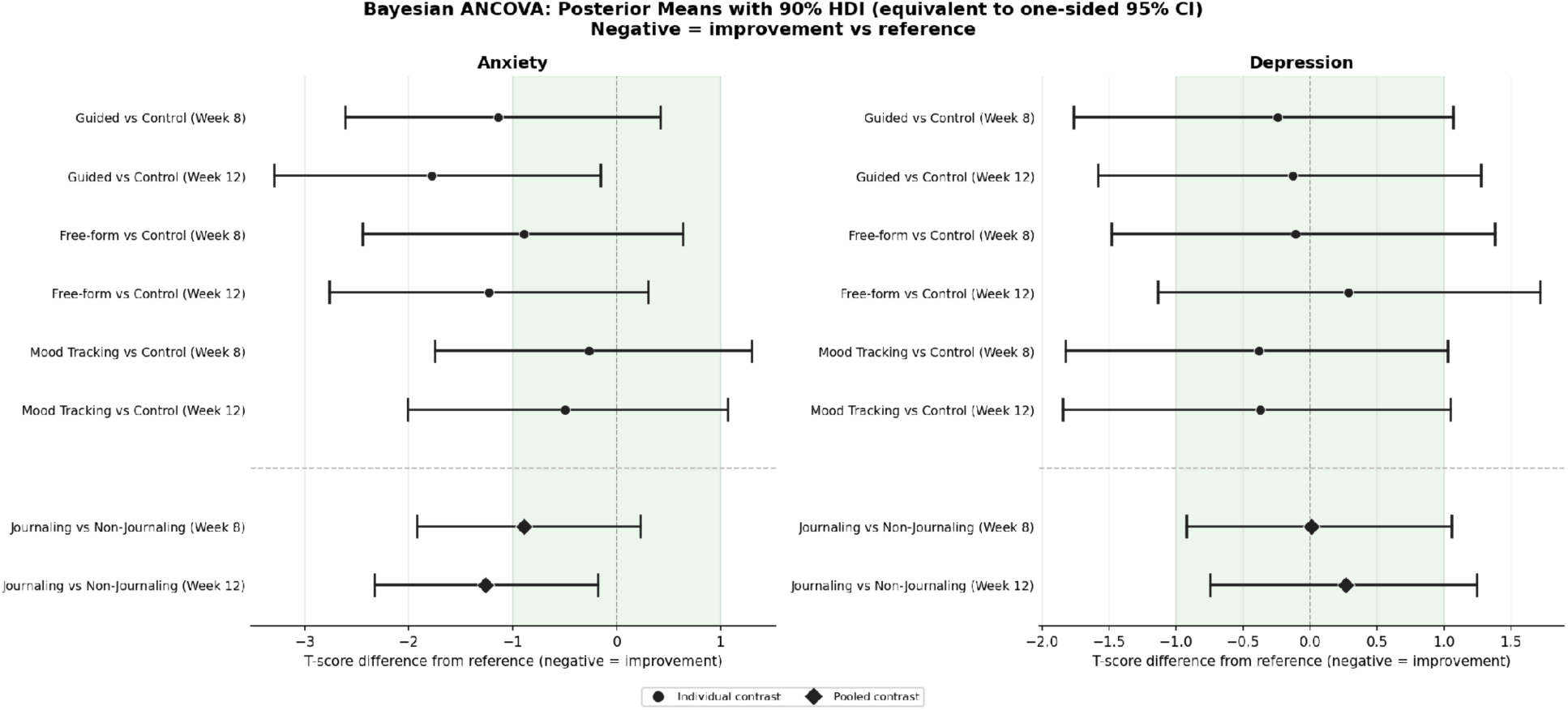
Bayesian ANCOVA estimates of intervention effects on anxiety and depression. Posterior mean differences in PROMIS T-scores are shown for each intervention arm relative to control at Week 8 and Week 12, along with pooled contrasts (journaling vs non-journaling). Points indicate posterior means and horizontal bars denote 90% highest density intervals (HDI), corresponding to directional (one-sided) inference. Negative values indicate symptom improvement relative to the reference group. Shaded regions denote the region of practical equivalence (ROPE; ±1 T-score point). Stronger evidence for symptom reduction observed for journaling conditions in anxiety at Week 12, and no meaningful effects observed for depression.

**Table S2.** Participant Demographics by Study Arm.

|  | **Guided (163)** | **Unguided (167)** | **Mood check (169)** | **Control (166)** | ***P*** |
|---|---|---|---|---|---|
| Age | 22.5 ± 2.0 | 22.3 ± 2.1 | 22.5 ± 2.0 | 22.2 ± 2.2 | .420 |
| Education | 15.2 ± 1.8 | 15.1 ± 1.8 | 15.1 ± 1.8 | 15.1 ± 1.7 | .905 |
| PROMIS Anxiety T | 59.3 ± 6.5 | 59.4 ± 6.8 | 58.5 ± 7.1 | 58.6 ± 7.6 | .518 |
| PROMIS Depression T | 58.3 ± 5.7 | 57.8 ± 6.3 | 58.4 ± 6.4 | 57.4 ± 6.6 | .403 |
| GAD-7 | 8.3 ± 3.2 | 8.7 ± 3.1 | 8.0 ± 3.2 | 8.1 ± 3.2 | .202 |
| PHQ-8 | 8.9 ± 3.1 | 8.2 ± 3.2 | 8.4 ± 3.3 | 8.5 ± 3.0 | .228 |
| WHODAS (raw total) | 71.9 ± 21.8 | 69.3 ± 21.3 | 69.7 ± 21.0 | 71.1 ± 22.9 | .667 |
| Female | 91 (55.8) | 106 (63.5) | 94 (55.6) | 107 (64.5) | .524 |
| White | 89 (54.6) | 98 (58.7) | 101 (59.8) | 92 (55.4) | .316 |

*Note.* The table summarizes the full assigned cohort: guided journaling plus mood check (n=163), unguided journaling plus mood check (n=167), mood check only (n=169), and passive control (n=166). Continuous variables are presented as mean ± SD; categorical variables as n (%). P values compare characteristics across the four assigned conditions.

### Supplementary Results: Sensitivity to minimum observation threshold

To assess whether the volatility findings depended on the requirement that users contribute at least 10 observations, we repeated the analysis across minimum-observation thresholds ranging from 8 to 24 observations. For these sensitivity analyses, users were required to have sufficient data to estimate both volatility and acute-drop rate, including at least five evaluable temporal windows.

At the primary ≥10-observation threshold, 289 users were eligible for volatility estimation and 253 had sufficient temporal windows for estimation of acute-drop rate. In this paired analytic sample, pleasantness volatility was strongly associated with valence-drop rate (r = .647, 95% CI [.566, .715], P < .001), while energy volatility showed a smaller cross-dimensional association (r = .389, 95% CI [.288, .492], P < .001).

Associations were highly stable as the minimum-observation requirement became more stringent. Pleasantness-volatility correlations ranged from .647 to .670 across thresholds, while energy-volatility correlations ranged from .387 to .433 (Table S3). Because the requirement of five evaluable windows itself imposes an effective minimum data-density requirement, thresholds from 8 through 16 observations yielded the same final paired analytic sample (n = 253). Results remained essentially unchanged when the requirement was increased to 20 observations (n = 238) or 24 observations (n = 224), indicating that the observed associations were not driven by participants with sparse longitudinal data.

**Table S3.**
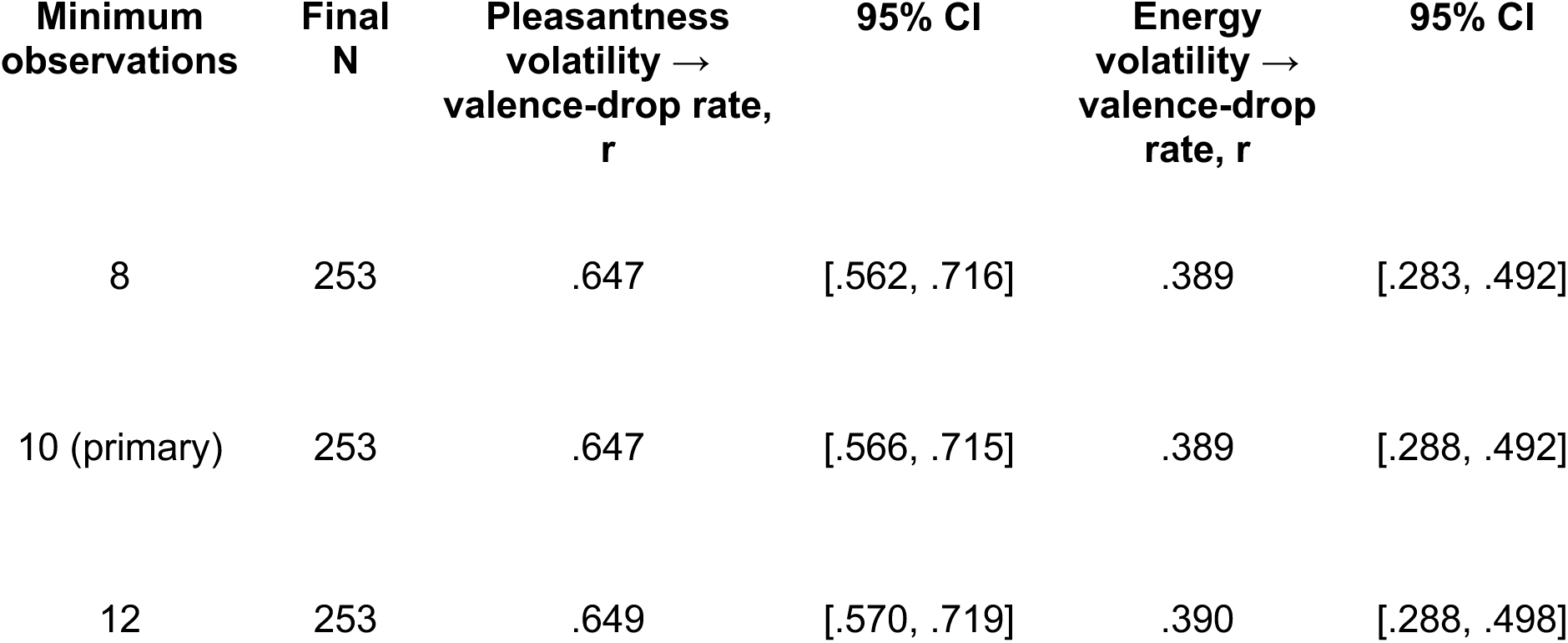
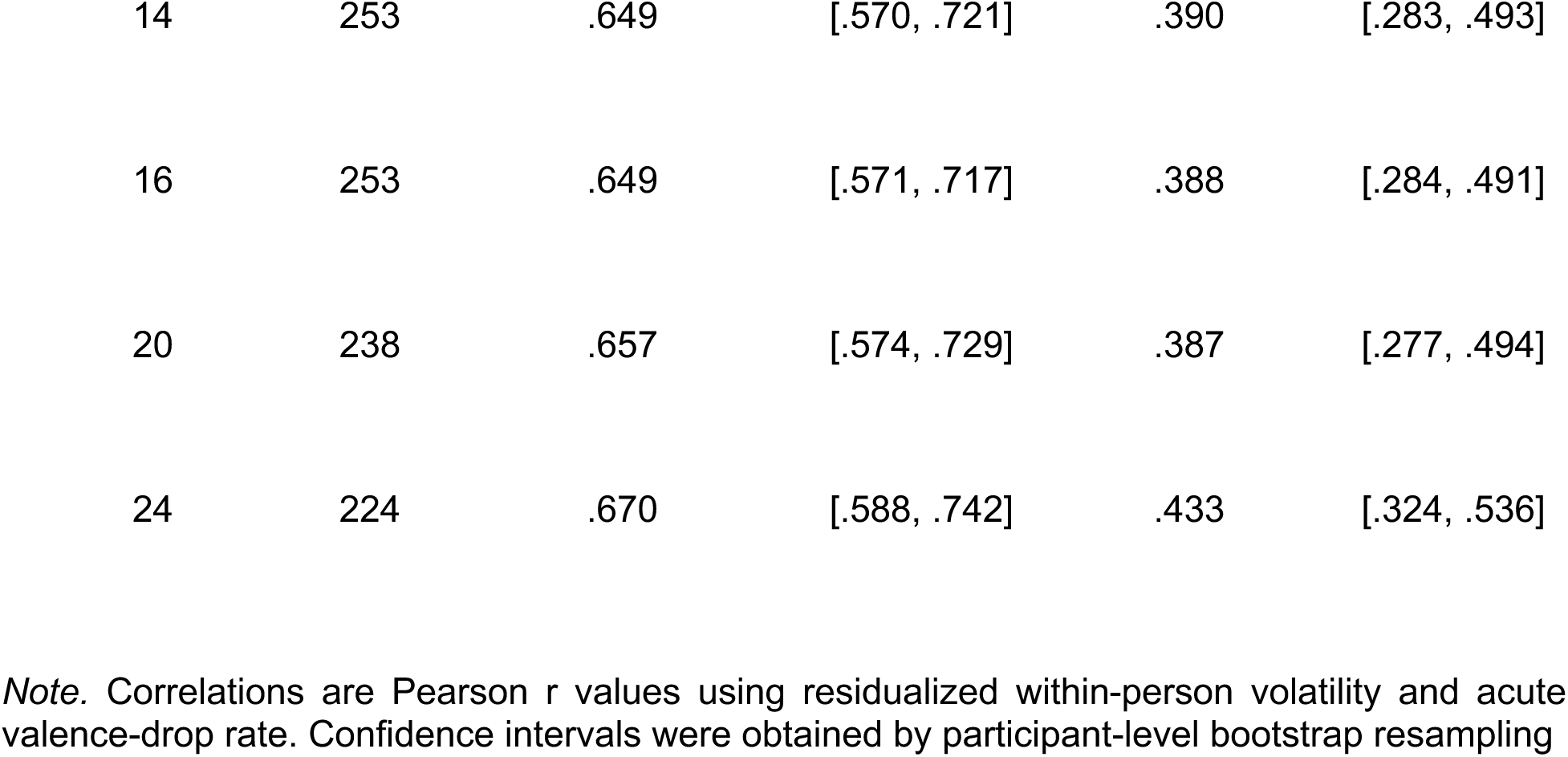
Sensitivity of volatility–drop associations to the minimum number of observations per user valence-drop rate. Confidence intervals were obtained by participant-level bootstrap resampling.

| **Minimum observations** | **Final N** | **Pleasantness volatility → valence-drop rate, r** | **95% CI** | **Energy volatility → valence-drop rate, r** | **95% CI** |
|---|---|---|---|---|---|
| 8 | 253 | .647 | [.562, .716] | .389 | [.283, .492] |
| 10 (primary) | 253 | .647 | [.566, .715] | .389 | [.288, .492] |
| 12 | 253 | .649 | [.570, .719] | .390 | [.288, .498] |
| 14 | 253 | .649 | [.570, .721] | .390 | [.283, .493] |
| 16 | 253 | .649 | [.571, .717] | .388 | [.284, .491] |
| 20 | 238 | .657 | [.574, .729] | .387 | [.277, .494] |
| 24 | 224 | .670 | [.588, .742] | .433 | [.324, .536] |

*Note.* Correlations are Pearson r values using residualized within-person volatility and acute valence-drop rate. Confidence intervals were obtained by participant-level bootstrap resampling

### Supplementary Results: Sensitivity of transition dynamics to the temporal linking window

To assess whether transition and persistence estimates depended on the primary ≤48-hour interval between successive entries, analyses were repeated using maximum inter-entry intervals of ≤24 and ≤72 hours. The number of eligible mood transitions was 5,964 from 305 users at ≤24 hours, 10,227 from 317 users at ≤48 hours, and 11,157 from 319 users at ≤72 hours. Because automated risk classification was available only in the unguided journaling arm, risk-transition analyses were restricted to that condition and to observations with evidence that automated classification had occurred. This yielded 1,405 eligible risk transitions from 94 users at ≤24 hours, 2,640 transitions from 99 users at ≤48 hours, and 2,927 transitions from 100 users at ≤72 hours.

Mood-state persistence was highly consistent across temporal specifications. Calm/Relaxed remained the most persistent state at each interval (42.3%, 41.5%, and 40.9% at ≤24, ≤48, and ≤72 hours, respectively), whereas Anxious/Agitated remained the least persistent (17.8%, 13.2%, and 11.6%). Happy/Alert and Sad/Withdrawn showed intermediate persistence across all three specifications.

Risk-state patterns were also broadly consistent across temporal specifications. Stable classifications showed high user-weighted persistence at each interval (88.6%, 88.9%, and 87.9% at ≤24, ≤48, and ≤72 hours, respectively). Concerning classifications showed lower persistence (20.0%, 12.4%, and 11.7%, respectively). High-Risk persistence estimates were less precise because relatively few users contributed eligible transitions from a High-Risk state (4, 8, and 9 users, respectively); corresponding persistence estimates were 0.0%, 9.4%, and 11.1%. Accordingly, High-Risk persistence estimates should be interpreted descriptively. Overall, the qualitative structure of mood and risk transitions was not dependent on the choice of temporal linking window.

We additionally repeated the transition analysis restricting successive observations to the same local calendar day. This yielded 1,181 mood transitions from 205 users (median inter-entry interval 8 hours, IQR 3–11) and 151 risk transitions from 44 users (median inter-entry interval 11 hours, IQR 5–15). The same broad persistence structure was preserved. Happy/Alert and Calm/Relaxed showed the greatest mood persistence (48.7% and 48.1%, respectively), whereas

Anxious/Agitated remained the least persistent state (18.4%). Stable risk classifications remained highly persistent (85.7%), while Concerning persistence was 21.2%. Only one user contributed a same-day transition from a High-Risk state, precluding meaningful estimation of High-Risk persistence. These findings indicate that the principal transition patterns were also evident when analyses were restricted to observations occurring within the same calendar day.

**Figure S2.**
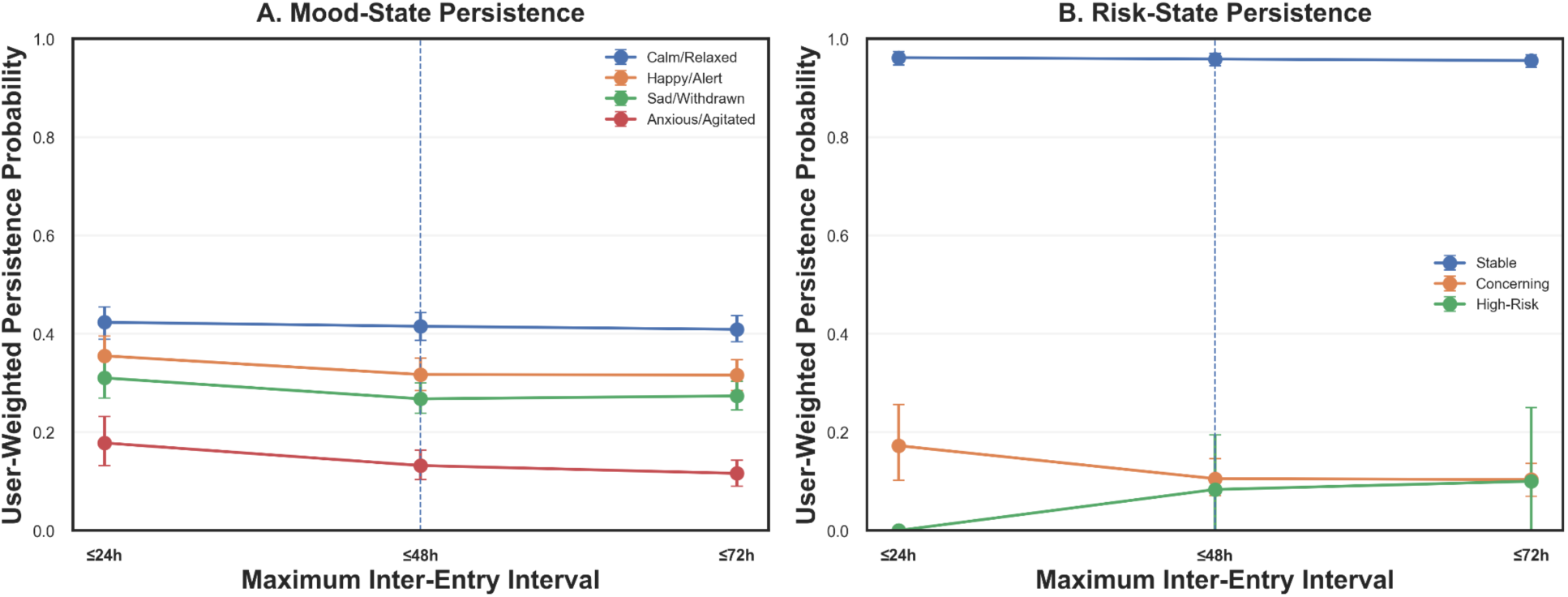
Sensitivity of state persistence to the temporal transition window. User-weighted self-transition probabilities for mood states (A) and risk states (B) using maximum inter-entry intervals of ≤24, ≤48, and ≤72 hours. Points represent mean user-level persistence probabilities and error bars indicate 95% participant-bootstrap confidence intervals. The ≤48-hour interval represents the primary analysis. Mood-state ordering was preserved across temporal specifications, while Stable and Concerning risk classifications showed broadly consistent patterns. High-Risk estimates were based on relatively few contributing users and should therefore be interpreted descriptively.

### Temporal-null analysis of volatility-drop associations

Because volatility and threshold-defined drop frequency are mathematically related, we evaluated whether the observed associations exceeded those expected from each participant’s marginal affective variability alone. Analyses were restricted to the 253 users with sufficient data to estimate both volatility and acute-drop rate, thereby avoiding structural assignment of zero drop rates to users with insufficient temporal windows. Within each user, daily pleasantness and energy values were jointly reordered using full shuffling and block-preserving permutations of 2, 5, and 7 observations, with random circular phase. These permutations preserved each participant’s marginal affect distribution and pleasantness–energy pairing while disrupting temporal sequence. Across 1,000 permutations per specification, the observed pleasantness association (r = .647) did not exceed the permutation distributions (null means = .640–.649; empirical Ps = .413–.557), nor did the energy association (r = .389; null means = .353–.361; empirical Ps = .176–.225). Thus, the strong participant-level association is largely attributable to marginal affective variability, with no evidence that temporal ordering contributes additional association.

### Prior sensitivity of the Bayesian models

The Bayesian models reported in the main text used weakly informative priors on the group and timepoint-by-group coefficients, Normal(0, 10). On the PROMIS T-score metric this implies a prior standard deviation of 14.1 points for the week-12 contrast, or roughly ±28 points at two standard deviations — a range wider than the clinically meaningful span of the instrument. Because such a prior is nearly flat relative to the likelihood, the resulting probability of direction closely approximates the one-sided frequentist P value and should not be interpreted as independent corroboration of the frequentist contrasts. In a post hoc analysis, we therefore refit the models across a ladder of priors on the treatment coefficients, holding the model specification, the analytic sample, and the intercept, timepoint, residual, and random-intercept priors fixed. For each outcome, arm-level contrasts were re-estimated in the four-arm model and the pooled contrast in the separately fitted pooled model, matching the corresponding analyses in the main text.

Three specifications were treated as candidate substantive beliefs: Normal(0, 1), Normal(0, 2), and Normal(0, 5) on the group coefficient, with interaction priors set to half the corresponding scale, since the interaction represents the change in treatment effect between week 8 and week 12 and should a priori be smaller than the effect itself. Given a within-sample SD of approximately 7 T-score points, these correspond to prior beliefs that effects larger than approximately d = 0.3, d = 0.6, and d = 1.4 are unlikely. Two further specifications, Normal(0, 10) — the primary analysis — and Normal(0, 20), were retained as diagnostics rather than as plausible beliefs; their close agreement quantifies how little information the primary prior contributed beyond the likelihood. For each specification we report the posterior mean, 90% HDI, pd, the probability that the effect exceeded the region of practical equivalence (P[β < −1]), and the proportion of posterior mass within the ROPE (±1 T-score point).

The direction of the anxiety estimate was stable across the ladder. For guided journaling at week 12, the posterior mean was −1.78 (P[effect < 0] = 0.97) under the primary specification and −1.79 (0.97) under a near-flat prior, confirming that the primary prior contributed little beyond the likelihood. Estimates attenuated monotonically as the prior was tightened: −1.70 (0.97) under Normal(0, 5), −1.38 (0.95) under Normal(0, 2), and −0.94 (0.92) under Normal(0, 1). The pooled journaling contrast followed the same pattern, from −1.27 to −0.91. Unguided journaling was less stable, with the directional probability falling from 0.90 under the primary specification to 0.80 under the most skeptical prior. Posterior magnitude quantities showed the same prior dependence. The probability that the guided-journaling effect exceeded the ROPE fell from 0.79 under the primary specification to 0.46 under Normal(0, 1), and for the pooled contrast from 0.65 to 0.44; under the most skeptical prior, 53% and 56% of posterior mass respectively fell within ±1 T-score point. Unlike the anxiety estimates, depression estimates were essentially invariant to prior specification, with substantial posterior mass within the specified ROPE. This describes the estimates under these models and does not establish absence of any clinically meaningful effect in the full assigned population.

**Table S4.** Sensitivity of estimates to the prior specification.

| Outcome | Contrast | Timepoint | N(0, 1) | N(0, 2) | N(0, 5) | N(0, 10) primary | N(0, 20) flat |
|---|---|---|---|---|---|---|---|
| Anxiety | Guided journaling | Week 12 | -0.94 (0.92; 53%) | -1.38 (0.95; 32%) | -1.70 (0.97; 23%) | -1.78 (0.97; 20%) | -1.79 (0.97; 20%) |
| Anxiety | Guided journaling | Week 8 | -0.72 (0.88; 67%) | -1.00 (0.90; 50%) | -1.11 (0.90; 43%) | -1.14 (0.89; 43%) | -1.12 (0.88; 43%) |
| Anxiety | Unguided journaling | Week 12 | -0.54 (0.80; 75%) | -0.92 (0.87; 53%) | -1.13 (0.89; 44%) | -1.23 (0.90; 39%) | -1.24 (0.90; 39%) |
| Anxiety | Unguided journaling | Week 8 | -0.47 (0.78; 80%) | -0.76 (0.84; 61%) | -0.86 (0.83; 55%) | -0.89 (0.83; 52%) | -0.88 (0.82; 52%) |
| Anxiety | Mood tracking | Week 12 | 0.05 (0.53; 86%) | -0.21 (0.60; 76%) | -0.41 (0.67; 67%) | -0.50 (0.71; 66%) | -0.49 (0.70; 65%) |
| Anxiety | Mood tracking | Week 8 | 0.04 (0.53; 90%) | -0.16 (0.58; 79%) | -0.22 (0.60; 72%) | -0.27 (0.61; 69%) | -0.24 (0.60; 71%) |
| Anxiety | Pooled journaling | Week 12 | -0.91 (0.95; 56%) | -1.14 (0.97; 42%) | -1.22 (0.97; 37%) | -1.26 (0.97; 35%) | -1.27 (0.98; 35%) |
| Anxiety | Pooled journaling | Week 8 | -0.72 (0.92; 71%) | -0.84 (0.92; 60%) | -0.86 (0.91; 57%) | -0.89 (0.91; 57%) | -0.89 (0.92; 56%) |
| Depression | Guided journaling | Week 12 | -0.12 (0.57; 88%) | -0.15 (0.58; 80%) | -0.13 (0.56; 75%) | -0.13 (0.56; 74%) | -0.15 (0.57; 74%) |
| Depression | Guided journaling | Week 8 | -0.10 (0.56; 91%) | -0.16 (0.59; 82%) | -0.20 (0.60; 76%) | -0.24 (0.62; 74%) | -0.24 (0.61; 74%) |
| Depression | Unguided journaling | Week 12 | 0.20 (0.38; 87%) | 0.25 (0.37; 78%) | 0.29 (0.36; 74%) | 0.29 (0.37; 72%) | 0.30 (0.37; 72%) |
| Depression | Unguided journaling | Week 8 | 0.09 (0.43; 91%) | 0.02 (0.49; 83%) | -0.06 (0.53; 76%) | -0.11 (0.55; 74%) | -0.09 (0.54; 76%) |
| Depression | Mood tracking | Week 12 | -0.28 (0.68; 85%) | -0.37 (0.68; 75%) | -0.38 (0.67; 72%) | -0.37 (0.67; 71%) | -0.39 (0.67; 69%) |
| Depression | Mood tracking | Week 8 | -0.22 (0.65; 89%) | -0.30 (0.65; 79%) | -0.37 (0.67; 73%) | -0.38 (0.66; 71%) | -0.37 (0.67; 72%) |
| Depression | Pooled journaling | Week 12 | 0.18 (0.36; 93%) | 0.23 (0.34; 90%) | 0.28 (0.32; 87%) | 0.27 (0.32; 87%) | 0.27 (0.32; 87%) |
| Depression | Pooled journaling | Week 8 | 0.07 (0.45; 96%) | 0.07 (0.45; 93%) | 0.03 (0.48; 91%) | 0.01 (0.50; 90%) | 0.02 (0.49; 91%) |

Note. Cells show the posterior mean difference in PROMIS T-score points with the posterior probability supporting the corresponding hypothesis and the percentage of posterior mass within the region of practical equivalence (ROPE, ±1 T-score point) in parentheses; negative values favor the intervention. Following the convention used throughout, the journaling contrasts were evaluated directionally and the reported probability is P(effect < 0), whereas the mood tracking-only contrast was evaluated non-directionally and the reported probability is the probability of direction, which is bounded at 0.5 to 1 by construction; this mirrors the one-tailed and two-tailed conventions used in the corresponding frequentist contrasts. Estimates are from baseline-adjusted Bayesian mixed-effects ANCOVA models in the contamination-corrected sample (n = 498). Arm-level contrasts are relative to passive control and are estimated in the four-arm model; the pooled journaling contrast is relative to non-journaling (mood tracking plus passive control) and is estimated in the separately fitted pooled model, matching the corresponding analyses in the main text. Each column refits those models with a different prior on the group and timepoint-by-group coefficients, holding the model specification, the analytic sample, and all other priors fixed. Normal(0, 1), Normal(0, 2), and Normal(0, 5) are substantively motivated specifications, corresponding to prior beliefs that effects exceeding approximately d = 0.3, 0.6, and 1.4 are unlikely; Normal(0, 10) is the primary specification and Normal(0, 20) a near-flat limiting case, both retained as diagnostics rather than as plausible prior beliefs. Interaction priors were set to half the group-coefficient scale on the substantive specifications and equal to it on the diagnostic specifications. The Normal(0, 10) column reproduces the estimates reported in the main text. These probabilities are reported descriptively rather than as significance criteria. All models converged (R-hat ≤ 1.01). Highest density intervals and the diffuse-baseline-prior comparison are in the deposited analysis output.

**Figure S3.**
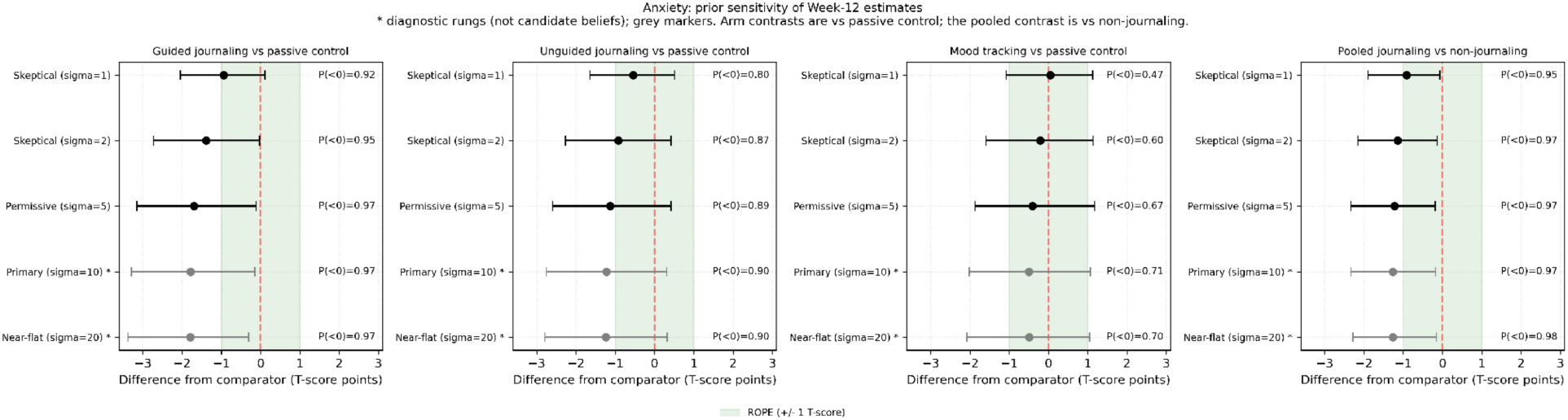
Prior sensitivity of week-12 anxiety estimates. Posterior mean differences at week 12 with 90% highest density intervals, shown across five prior specifications for guided journaling, unguided journaling, mood tracking, and the pooled journaling contrast. Arm-level contrasts are relative to passive control and are estimated in the four-arm model; the pooled journaling contrast is relative to non-journaling (mood tracking plus passive control) and is estimated in the separately fitted pooled model. Negative values favor the intervention. Shaded band indicates the region of practical equivalence (±1 T-score point); dashed line indicates no difference. The posterior probability supporting each hypothesis is printed beside each estimate: P(effect < 0) for the journaling contrasts, which were evaluated directionally, and the probability of direction for mood tracking, which was not. Substantive prior specifications are shown in black; diagnostic specifications — the primary analysis, Normal(0, 10), and a near-flat Normal(0, 20) — are shown in grey and marked with an asterisk. Estimates attenuate as the prior tightens: under the most skeptical specification all three journaling estimates fall within the ROPE (guided −0.94, unguided −0.54, pooled −0.91), with 53–75% of posterior mass inside it. The direction of all three is nonetheless preserved across every specification examined.

**Figure S4.**
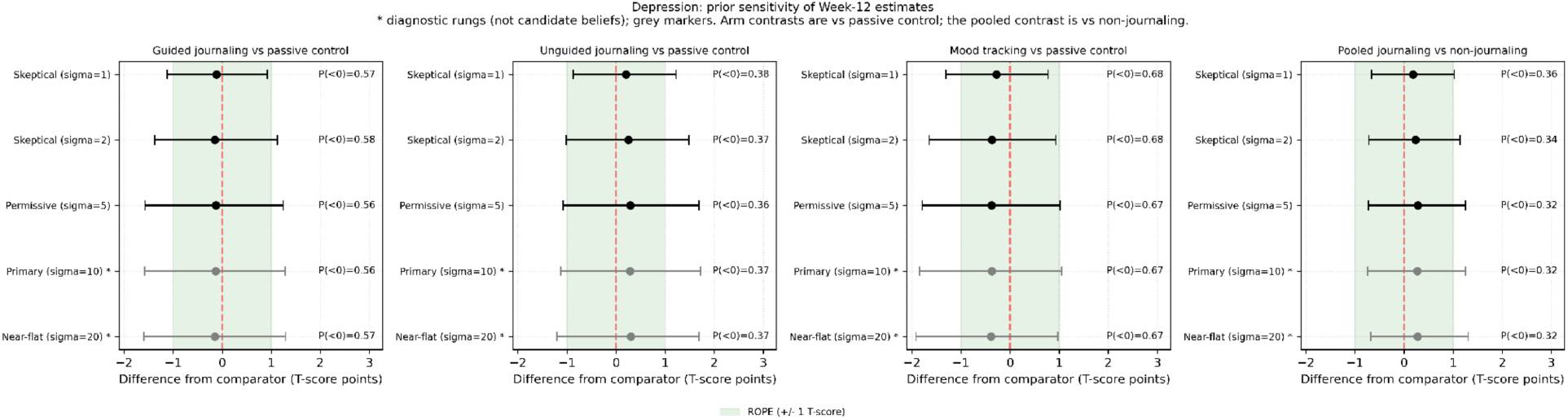
Prior sensitivity of week-12 depression estimates. Panels, prior specifications, and annotation follow Figure S3. Estimates remained centered near zero across all prior specifications, ranging from −0.39 to +0.30 T-score points, with 69–93% of posterior mass within the ROPE. Depression estimates were essentially unchanged by prior specification; the sign differed across contrasts and every estimate fell well within the region of practical equivalence.

